# Impact of Preexisting Rare Diseases on COVID-19 Severity, Reinfection, and Long COVID, and the Modifying Effects of Vaccination and Antiviral Therapy: A Retrospective Study from the N3C Data Enclave

**DOI:** 10.1101/2025.07.09.25331138

**Authors:** Arjun S. Yadaw, David K Sahner, Eric Sid, Emily Y. Chew, Dominique Pichard, Ewy A. Mathé, the N3C Consortium

## Abstract

**Background:** Over 10,000 rare diseases (RDs) affect more than 300 million people globally, yet their influence on COVID-19 severity, reinfection risk, and long COVID remains poorly understood. This study evaluates the impact of RDs on these outcomes and examines the effectiveness of vaccination and antiviral treatments among individuals with and without RDs.

**Methods:** We conducted a retrospective cohort study using harmonized electronic health records (EHRs) from the National COVID Cohort Collaborative (N3C), encompassing 21,704,702 individuals, including 4,825,605 with confirmed SARS-CoV-2 infection between Jan 1, 2020, and Jan 4, 2024. RDs were defined using 12,003 conditions curated from GARD and Orphanet, mapped to OMOP concepts, and classified into 18 RD classes based on medical specialty involvement. Primary outcomes included: (1) COVID-19 severity (hospitalization and life-threatening disease), (2) long COVID, and (3) SARS-CoV-2 reinfection. We applied multivariable logistic regression with inverse probability of treatment weighting and reported adjusted odds ratios with 95% confidence intervals and associated p-values. Models were controlled for demographics, comorbidities, and exposure to vaccination and antiviral treatments.

**Findings:** Of 21,704,702 individuals, we identify 4,825,605 COVID-19 positive individuals, 6.36% had RDs, with markedly higher rates of rare disease (RD) patients that have life-threatening illness (16% vs. 6.1% without life-threatening illness) and that are hospitalized (13% vs. 6.0% without hospitalization). Otorhinolaryngologic diseases showed the highest risk of life-threatening outcomes (OR 4.51; 95% CI 3.81-5.33), followed by developmental defect during embryogenesis (OR 1.84; 95% CI 1.72-1.98) and cardiac conditions (OR 1.79; 95% CI 1.51-2.11). Hospitalization risk was highest for otorhinolaryngologic (OR 2.90; 95% CI 2.61-3.23), developmental defect during embryogenesis (OR 2.06; 95% CI 1.97-2.16), and hematologic and endocrine diseases (OR 1.81; 95% CI 1.75-1.87 and OR 1.81; 95% CI 1.64-1.99, respectively).

In patients with RDs, vaccination alone or antiviral treatment alone was associated with reduced odds of life-threatening COVID-19 disease compared to non-vaccinated individuals (OR 0.71; 95% CI 0.66-0.77 and OR 0.33; 95% CI 0.26-0.42, respectively). The combination of both vaccination and antiviral treatment showed the greatest reduction in odds ratio (OR 0.24; 95% CI 0.20-0.27). Similar results were observed in patients without RDs. In contrast, vaccination or antiviral therapy alone, compared to no intervention, did not significantly reduce long COVID risk in RD patients, although these interventions alone did result in a lower odds ratio in patients without RD. However, their combination was protective in both groups. Vaccination alone, compared to no vaccination, also reduced the risk of reinfection across RD and non-RD populations.

**Interpretation:** RD patients face elevated risks of severe COVID-19 outcomes. While vaccination and antivirals significantly reduce the acute severity of illness, their impact on long COVID appears limited in this population. Notably, vaccination was protective against COVID-19 reinfection in both RD and non-RD populations. These findings highlight the need for targeted strategies to protect RD patients beyond current interventions, particularly in preventing long-term complications.

**Funding:** This work was supported in part by the intramural and extramural programs at NCATS (ZIA ZICTR000410).

## Introduction

Globally, over 777 million individuals have been infected by the SARS-CoV-2 virus as of 20^th^ April 2025^1^. While much attention has been given to the acute phase of the infection, emerging evidence suggests that the long-term consequences of COVID-19, often referred to as "long COVID," and the risk of reinfection are significant public health concerns^2–4^. Numerous studies have shown that a significant proportion of individuals with COVID-19 continue to experience persistent symptoms lasting beyond four weeks, with some cases extending for several months or even years after the initial infection^5–7^. RDs, defined in the United States (US) as conditions affecting fewer than 200,000 individuals in the US, encompass over 10,000 distinct disorders that collectively impact more than 30 million people^8–10^. Individuals with RDs represent a unique subset due to their distinct and heterogenous pathophysiological profiles and often compromised immune systems^11^.

Despite growing interest, significant gaps remain in understanding the impact of RDs on COVID-19 severity. A recent study has indicated that patients with underlying health conditions, including RDs, are at a higher risk for severe COVID-19 outcomes, such as mortality^12^. While these findings underscore the relevance of RDs in the context of the pandemic, the scope of prior research remains narrow. Notably, a population-wide study in England utilizing linked electronic health record (EHR) data from over 58 million individuals examined the prevalence of only 331 RDs and their association with COVID-19-related mortality^13^. This highlights that, despite the breadth of available data, evaluations of COVID-19 outcomes have thus far been limited to a small subset of the more than 10,000 known RDs, leaving a substantial knowledge gap.

One key challenge in evaluating the broader RD population is the heterogeneity in disease etiology, as well as variation in diagnostic and treatment practices. To help address classification issues and ensure that each RD is assigned to a single category, Orphanet has developed RD linearization to facilitate evaluation of RD groups by medical specialty^14^. A recent investigation further advanced this effort by mapping a larger number of RDs to diagnosis codes (SNOMED-CT and ICD-10) used in the EHR and assessing the risk ratio of COVID-19 severity across RD classes based on medical specialty^15^. Although progress has been made, the risks and burden associated with preexisting RDs in relation to long COVID and reinfection remain poorly understood. There is a critical need to evaluate these outcomes and assess the effects of prevention and intervention strategies such as vaccination and antiviral treatments to better inform public health and clinical decision-making.

In this retrospective study, we leveraged harmonized EHR data from the National COVID Cohort Collaborative (N3C), the largest integrated COVID-19 dataset in the United States, to evaluate the impact of preexisting RDs on COVID-19 severity, long COVID, and reinfection. We also examined how vaccination and Food and Drug Administration (FDA) approval and Emergency Use Authorization (EUA) of antiviral treatments such as Paxlovid, and Molnupiravir affect these outcomes in individuals with and without RDs. Vaccination has been shown to significantly reduce the severity of COVID-19 and the risk of hospitalization^16^, while antivirals have demonstrated efficacy in lowering hospitalization and mortality rates in high-risk patients^17,18^. This study addresses four critical questions: (i) Do individuals with specific RD classes or RD as a group face greater risk of severe COVID-19 outcomes (hospitalization, life-threatening)? (ii) Are RD patients more susceptible to developing long COVID? (iii) Is the risk of reinfection elevated in this RD population? (iv) How do vaccination and antiviral therapies modify these risks across rare and non-rare disease groups? By addressing these questions, our study aims to fill key evidence gaps and inform clinical decision-making and policy development for a medically underserved RD population.

## Methods

### Study design and participants

We conducted a retrospective cohort study using harmonized EHR data from the N3C Enclave, the largest integrated COVID-19 EHR dataset in the United States^19^. The dataset aggregates clinical data from 83 participating institutions using the Observational Medical Outcomes Partnership (OMOP) Common Data Model within a secure Palantir Foundry environment. The study was approved by the Institutional Review Board (IRB) under the N3C data use agreement [IRB00249128]. The N3C enclave released version V154 from Limited Datasets data (N=21,704,702 individuals) was used and includes individuals entered at or before January 4th, 2024. We identified a subset of 4,825,605 individuals who tested positive for COVID-19 via RT-PCR, antigen (Ag), or clinical diagnosis between January 1, 2020, and January 4, 2024 (**Figure 1**). We excluded individuals with missing age or sex, those under 1 year of age, individuals with fewer than one clinical encounter before or after the COVID-19 diagnosis date, and those from sites failing data quality checks. Historical data on comorbidities, medication exposures, and other clinical variables were available as early as January 1, 2018^19^.

**Figure 1:**
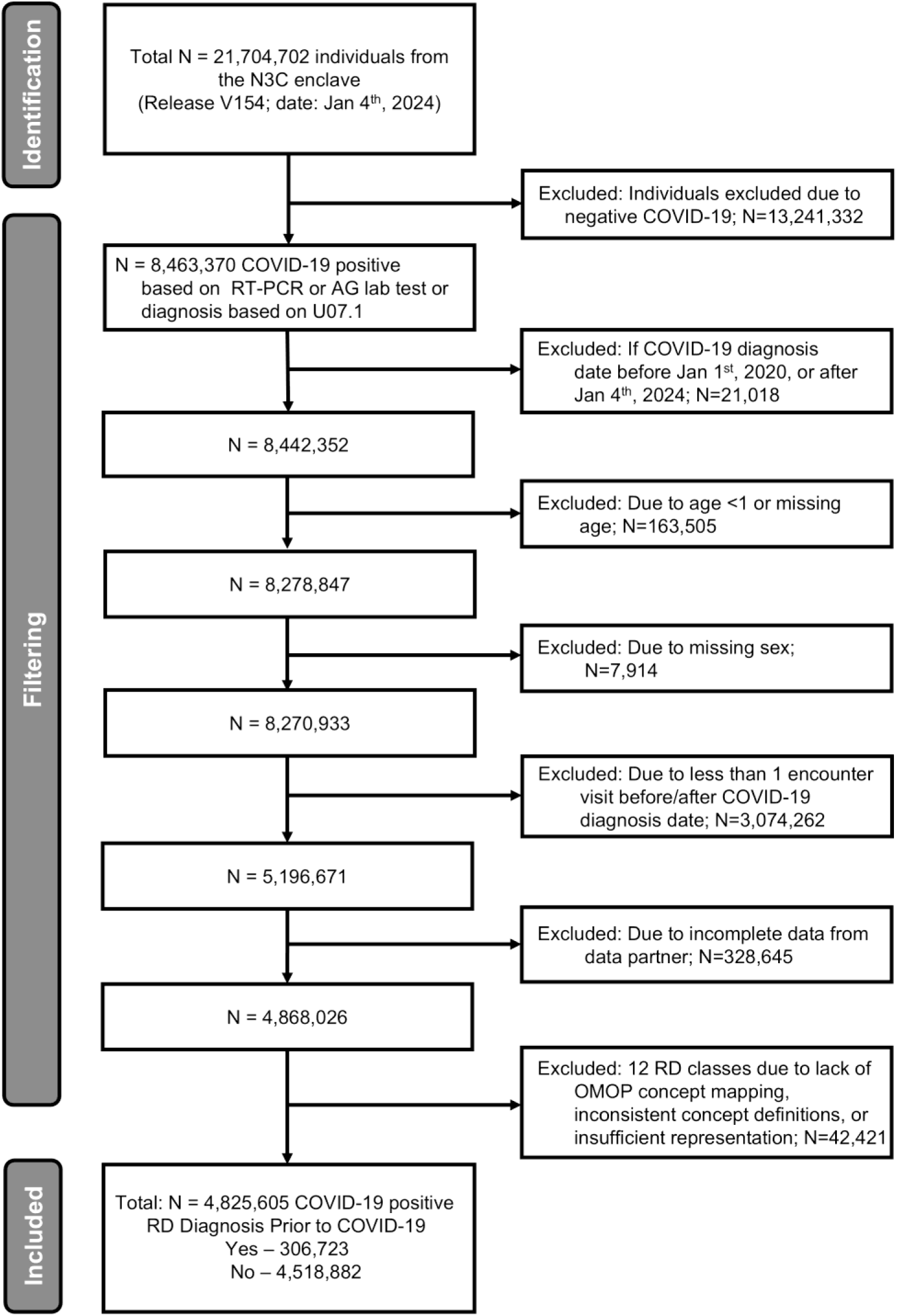
Workflow to define the RD patients within N3C. EHR data on 21,704,702 patients were available in the N3C enclave release version V154. Of these, 4,825,605 were defined as COVID-19 positive between Jan 1^st^, 2020, or after Jan 4^th^, 2024 (see **Methods**). These patients were further filtered for: non missing values for age and sex, at least one encounter visit before and after COVID-19 diagnosis date, incomplete data from data partners, inconsistent mappings or insufficient representation. Lastly, patients with COVID-19 were stratified into two groups, those with and without RDs.

### Rare disease diagnosis definitions

We curated 12,003 unique RDs from the Genetic and Rare Disease Information Center (GARD) and mapped them to SNOMED-CT and ICD-10 codes using Orphanet (July 2023 version), which employs the OHDSI Atlas (OMOP data model)^15^. To increase coverage of RDs, we included descendant codes of the resulting RD SNOMED-CT codes using the OHDSI Atlas tool^20^. We next excluded group of disorders (e.g. disorder of carbohydrate metabolism brings type II diabetes and many more descendants which are not RDs) and phenotypes, as defined by the Human Phenotype Ontology (HPO) via HPO API^21^. The finalized RD-specific SNOMED-CT and ICD-10 codes were converted to OMOP concept IDs using the N3C concept table. We applied these concept IDs to identify COVID-19-infected individuals with a prior RD diagnosis. When the resulting incidence exceeded six per ten thousand (reflecting the United States Definition of RDs), manual review by a clinician was performed, resulting in removal of 26 non-rare diseases^15^.

RDs were classified into 30 classes based on the Orphanet linearization classification based on the Orphanet Linearization system^14^. Twelve classes were excluded from the analysis due to lack of OMOP concept mapping, inconsistent concept definitions, or insufficient representation^15^. The excluded classes are abdominal surgical, circulatory system, disorder due to toxic effects, genetic, gynecologic or obstetric, infertility, maxillofacial surgical, odontologic, surgical cardiac, surgical thoracic, transplantation, and urogenital. Ultimately, eighteen rare disease classes (rare bone, cardiac, developmental, endocrine, gastroenterologic, hematologic, hepatic, immune, inborn errors of metabolism, infectious, neoplastic, neurologic, ophthalmic, otorhinolaryngologic, renal, respiratory, skin, and systemic or rheumatologic diseases) were considered in the study. We note that an RD can only belong to a single Orphanet linearization class, where the priority of the assigned class for RDs with multiple potential classes reflects the most severely affected body system, impact on prognosis, and primary specialist. The final cohort comprises 306,723 patients diagnosed with pre-existing RDs belonging to one of the18 RD classes listed above.

### Outcomes

The outcomes we evaluated were as follows:

1. COVID-19 severity: COVID-19 severity outcomes in N3C are based on the WHO Clinical Progression Scale^22^. We categorized binary outcomes as: (i) life-threatening disease (e.g., invasive mechanical ventilation, extracorporeal membrane oxygenation [ECMO], death)^23^, and (ii) hospitalization (yes/no) if the individual was hospitalized within 16 days of COVID-19 infection index date.
2. Long COVID: The CDC introduced the ICD-10 code U09.9 (Post COVID-19 condition, unspecified) on June 30, 2021, with implementation starting October 1, 2021. Due to underreporting of long COVID (U09.9) in N3C, we additionally identified potential long COVID patients using a machine learning (RECOVER ML) model^2^, ICD-10 code B94.8, or observation concept ID 2004207791. The RECOVER xgboost ML model (≥0.75 probability) in the N3C Knowledge Store (release version Feb 28^th^, 2025) was used. Patients were included in the long COVID analysis if they had at least 90 days of follow-up after their initial COVID-19 diagnosis. Individuals who died within the first 90 days were excluded, but since death was comparatively infrequent, it is unlikely to have represented a competing risk.
3. COVID reinfection: COVID-19 reinfection is defined as a new positive SARS-CoV-2 PCR, antigen, or diagnosis occurring ≥60 days after the initial infection index date. The reinfection index date is the date of the new positive test. Subsequent reinfections follow the same 60-day rule. Although many studies use a 90-day threshold, evidence shows most individuals stop shedding virus within 60 days, supporting the 60-day reinfection definition^24,25^. Individuals were included in the reinfection analysis if they had at least 60 days of follow-up after their initial infection; those who died within this period were excluded. Since vaccination began in late 2020 in the US, we selected the cohort for reinfection analysis from January 1, 2021, to January 4, 2024.

### Covariates

In addition to the 18 RD class indicators described previously, we evaluated a set of demographic and clinical covariates as potential confounders. These included age (categorized as 1-20, 21-40, 41-65, and >65 years), body mass index (BMI: underweight <18.5, normal 18.5-25, overweight 25-30, obese ≥30 kg/m²), sex (male, female), race (Asian, Black or African American, White, unknown/missing), ethnicity (Hispanic or Latino, Non-Hispanic or Latino, unknown/missing), and smoking status (current or former smoker, non-smoker). Additional clinical covariates comprising preexisting comorbidities (diagnosis prior to that of COVID-19): cancer, cardiomyopathies, cerebrovascular disease, chronic lung disease, prior dementia, depression, diabetes, heart failure, HIV infection, hypertension, kidney disease, liver disease, coronary artery disease, myocardial infarction, peripheral vascular disease, and rheumatologic disease. To address multicollinearity, comorbidities closely related to each RD class were excluded from the corresponding models. For the rare cardiac disease models, cardiomyopathies, coronary artery disease, heart failure, and myocardial infarction were excluded. For the rare immune disease models, cancer, HIV infection, diabetes, and rheumatologic disease were excluded. For the rare renal disease models, kidney disease was excluded. For rare systemic or rheumatologic disease, we removed rheumatologic disease, HIV infection, diabetes, and rheumatologic disease. Also, antiviral treatments and vaccination exposures (described below) were considered in model building for different outcomes.

### Definitions of exposure to antivirals and vaccination

Antiviral treatment and vaccination exposure were considered, focusing on the Omicron variant period, as antiviral therapies became available under FDA approval and emergency use authorization (EUA) in late December 2021.

- Antiviral exposure was defined as receiving at least one dose of an oral antiviral, Paxlovid (nirmatrelvir/ritonavir) or Lagevrio (Molnupiravir) within 5 days of COVID-19 diagnosis.
- Vaccination status was defined as fully vaccinated if the patient had completed a primary vaccine series as defined by CDC at least 14 days prior to the initial COVID-19 diagnosis^26^.

### Statistical Analysis

We applied both univariate and multivariable logistic regression models to assess the association of each RD class with COVID-19 severity, adjusting for demographic and comorbidities covariables as described above. To evaluate the effects of vaccination and antiviral treatment among individuals with preexisting RDs compared to those without, we grouped all RD classes into a single "rare disease" group due to limited sample sizes within individual RD classes. To address baseline differences across treatment groups defined by vaccination status and/or antiviral use, we applied inverse probability of treatment weighting (IPTW) based on propensity scores, estimating the average treatment effect (ATE)^27^ for each model. Propensity scores were calculated using logistic regression models that included demographic and comorbidities variables. Stabilized weights were used to enhance robustness, and extreme weights were trimmed at the 99th percentile. Covariate balance between treatment groups was evaluated using standardized mean differences (SMD), with values below 0.1 indicating adequate balance. Finally, we applied robust logistic regression models incorporating both the trimmed IPTW weights and covariates to estimate adjusted odds ratios (ORs), 95% confidence intervals (CI), and p-values for the outcomes of interest. For models evaluating associations between RD class and outcomes, odds ratios with Bonferroni-adjusted p-values < 0.0028 were considered statistically significant. For models evaluations associations of vaccination, antiviral treatment, or both, odds ratios with Bonferroni-adjusted p-values < 0.017 are considered statistically significant. Statistical modeling (**Figure S1**) was conducted within the N3C enclave using SQL, Python (3.10.16), statsmodels (0.14.4), Patsy (1.0.1), Numpy (1.26.4), Pandas (1.5.3), Spark SQL (3.4.1.34), R libraries (WeightIt(1.4.0)), (cobalt(4.6.0)), (ggplot2(3.5.2)), (ggpubr(0.6.0)), (survey(3.8_3)), (dplyr(1.1.4)), (broom (1.0.8)), (gtsummary (2.1.0), tidyverse (2.0.0) & gt (1.0.0) libraries)^28^.

## Results

Using the N3C enclave, we analyzed EHR data from 4,825,605 individuals who tested positive for COVID-19 through Reverse Transcription Polymerase Chain Reaction (RT-PCR), Antigen (Ag), or clinical diagnosis between January 1, 2020, and January 4, 2024. Among these COVID-19 positive cases, 306,723 (6.36%) had RDs, 254,728 (5.28%) were hospitalized, and 113,510 (2.35%) had a life-threatening (or fatal) disease. Among patients who experienced life-threatening outcomes, a high proportion had underlying comorbidities, including hypertension (82,396; 73%), chronic lung disease (47,425; 42%), and diabetes (45,053; 40%). These same comorbidities were also prevalent among hospitalized patients, indicating a consistent pattern of increased severity among patients with these conditions (**Table 1**).

**Table 1:**
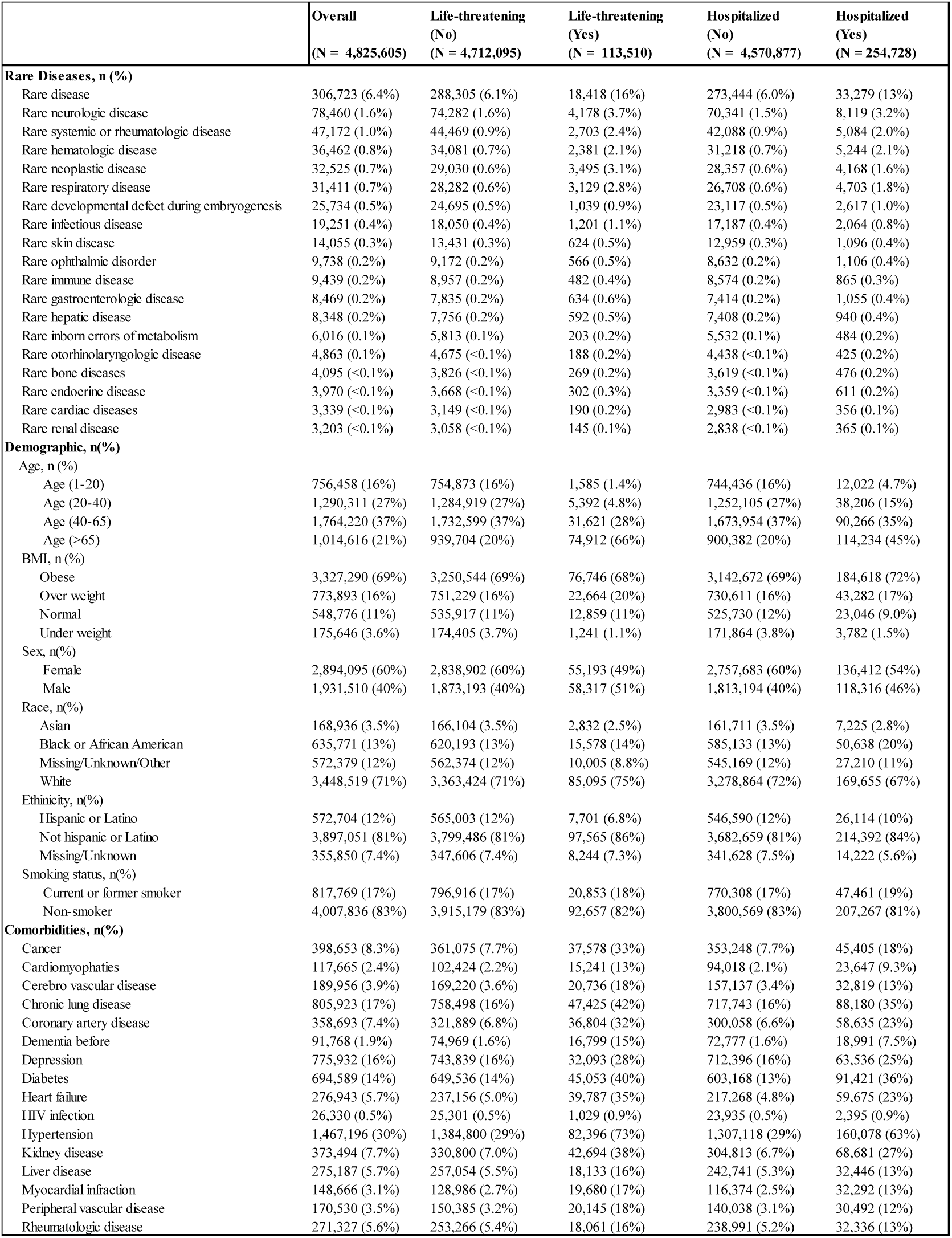
Characteristics of COVID-19 positive patients with and without rare disease.

|  | Overall<br>(N = 4,825,605) | Life-threatening<br>(No)<br>(N = 4,712,095) | Life-threatening<br>(Yes)<br>(N = 113,510) | Hospitalized<br>(No)<br>(N = 4,570,877) | Hospitalized<br>(Yes)<br>(N = 254,728) |
| --- | --- | --- | --- | --- | --- |
| **Rare Diseases, n (%)** |  |  |  |  |  |
| Rare disease | 306,723 (6.4%) | 288,305 (6.1%) | 18,418 (16%) | 273,444 (6.0%) | 33,279 (13%) |
| Rare neurologic disease | 78,460 (1.6%) | 74,282 (1.6%) | 4,178 (3.7%) | 70,341 (1.5%) | 8,119 (3.2%) |
| Rare systemic or rheumatologic disease | 47,172 (1.0%) | 44,469 (0.9%) | 2,703 (2.4%) | 42,088 (0.9%) | 5,084 (2.0%) |
| Rare hematologic disease | 36,462 (0.8%) | 34,081 (0.7%) | 2,381 (2.1%) | 31,218 (0.7%) | 5,244 (2.1%) |
| Rare neoplastic disease | 32,525 (0.7%) | 29,030 (0.6%) | 3,495 (3.1%) | 28,357 (0.6%) | 4,168 (1.6%) |
| Rare respiratory disease | 31,411 (0.7%) | 28,282 (0.6%) | 3,129 (2.8%) | 26,708 (0.6%) | 4,703 (1.8%) |
| Rare developmental defect during embryogenesis | 25,734 (0.5%) | 24,695 (0.5%) | 1,039 (0.9%) | 23,117 (0.5%) | 2,617 (1.0%) |
| Rare infectious disease | 19,251 (0.4%) | 18,050 (0.4%) | 1,201 (1.1%) | 17,187 (0.4%) | 2,064 (0.8%) |
| Rare skin disease | 14,055 (0.3%) | 13,431 (0.3%) | 624 (0.5%) | 12,959 (0.3%) | 1,096 (0.4%) |
| Rare ophthalmic disorder | 9,738 (0.2%) | 9,172 (0.2%) | 566 (0.5%) | 8,632 (0.2%) | 1,106 (0.4%) |
| Rare immune disease | 9,439 (0.2%) | 8,957 (0.2%) | 482 (0.4%) | 8,574 (0.2%) | 865 (0.3%) |
| Rare gastroenterologic disease | 8,469 (0.2%) | 7,835 (0.2%) | 634 (0.6%) | 7,414 (0.2%) | 1,055 (0.4%) |
| Rare hepatic disease | 8,348 (0.2%) | 7,756 (0.2%) | 592 (0.5%) | 7,408 (0.2%) | 940 (0.4%) |
| Rare inborn errors of metabolism | 6,016 (0.1%) | 5,813 (0.1%) | 203 (0.2%) | 5,532 (0.1%) | 484 (0.2%) |
| Rare otorhinolaryngologic disease | 4,863 (0.1%) | 4,675 (<0.1%) | 188 (0.2%) | 4,438 (<0.1%) | 425 (0.2%) |
| Rare bone diseases | 4,095 (<0.1%) | 3,826 (<0.1%) | 269 (0.2%) | 3,619 (<0.1%) | 476 (0.2%) |
| Rare endocrine disease | 3,970 (<0.1%) | 3,668 (<0.1%) | 302 (0.3%) | 3,359 (<0.1%) | 611 (0.2%) |
| Rare cardiac diseases | 3,339 (<0.1%) | 3,149 (<0.1%) | 190 (0.2%) | 2,983 (<0.1%) | 356 (0.1%) |
| Rare renal disease | 3,203 (<0.1%) | 3,058 (<0.1%) | 145 (0.1%) | 2,838 (<0.1%) | 365 (0.1%) |
| **Demographic, n(%)** |  |  |  |  |  |
| Age, n (%) |  |  |  |  |  |
| Age (1-20) | 756,458 (16%) | 754,873 (16%) | 1,585 (1.4%) | 744,436 (16%) | 12,022 (4.7%) |
| Age (20-40) | 1,290,311 (27%) | 1,284,919 (27%) | 5,392 (4.8%) | 1,252,105 (27%) | 38,206 (15%) |
| Age (40-65) | 1,764,220 (37%) | 1,732,599 (37%) | 31,621 (28%) | 1,673,954 (37%) | 90,266 (35%) |
| Age (>65) | 1,014,616 (21%) | 939,704 (20%) | 74,912 (66%) | 900,382 (20%) | 114,234 (45%) |
| BMI, n (%) |  |  |  |  |  |
| Obese | 3,327,290 (69%) | 3,250,544 (69%) | 76,746 (68%) | 3,142,672 (69%) | 184,618 (72%) |
| Over weight | 773,893 (16%) | 751,229 (16%) | 22,664 (20%) | 730,611 (16%) | 43,282 (17%) |
| Normal | 548,776 (11%) | 535,917 (11%) | 12,859 (11%) | 525,730 (12%) | 23,046 (9.0%) |
| Under weight | 175,646 (3.6%) | 174,405 (3.7%) | 1,241 (1.1%) | 171,864 (3.8%) | 3,782 (1.5%) |
| Sex, n(%) |  |  |  |  |  |
| Female | 2,894,095 (60%) | 2,838,902 (60%) | 55,193 (49%) | 2,757,683 (60%) | 136,412 (54%) |
| Male | 1,931,510 (40%) | 1,873,193 (40%) | 58,317 (51%) | 1,813,194 (40%) | 118,316 (46%) |
| Race, n(%) |  |  |  |  |  |
| Asian | 168,936 (3.5%) | 166,104 (3.5%) | 2,832 (2.5%) | 161,711 (3.5%) | 7,225 (2.8%) |
| Black or African American | 635,771 (13%) | 620,193 (13%) | 15,578 (14%) | 585,133 (13%) | 50,638 (20%) |
| Missing/Unknown/Other | 572,379 (12%) | 562,374 (12%) | 10,005 (8.8%) | 545,169 (12%) | 27,210 (11%) |
| White | 3,448,519 (71%) | 3,363,424 (71%) | 85,095 (75%) | 3,278,864 (72%) | 169,655 (67%) |
| Ethnicity, n(%) |  |  |  |  |  |
| Hispanic or Latino | 572,704 (12%) | 565,003 (12%) | 7,701 (6.8%) | 546,590 (12%) | 26,114 (10%) |
| Not hispanic or Latino | 3,897,051 (81%) | 3,799,486 (81%) | 97,565 (86%) | 3,682,659 (81%) | 214,392 (84%) |
| Missing/Unknown | 355,850 (7.4%) | 347,606 (7.4%) | 8,244 (7.3%) | 341,628 (7.5%) | 14,222 (5.6%) |
| Smoking status, n(%) |  |  |  |  |  |
| Current or former smoker | 817,769 (17%) | 796,916 (17%) | 20,853 (18%) | 770,308 (17%) | 47,461 (19%) |
| Non-smoker | 4,007,836 (83%) | 3,915,179 (83%) | 92,657 (82%) | 3,800,569 (83%) | 207,267 (81%) |
| **Comorbidities, n(%)** |  |  |  |  |  |
| Cancer | 398,653 (8.3%) | 361,075 (7.7%) | 37,578 (33%) | 353,248 (7.7%) | 45,405 (18%) |
| Cardiomyopathies | 117,665 (2.4%) | 102,424 (2.2%) | 15,241 (13%) | 94,018 (2.1%) | 23,647 (9.3%) |
| Cerebro vascular disease | 189,956 (3.9%) | 169,220 (3.6%) | 20,736 (18%) | 157,137 (3.4%) | 32,819 (13%) |
| Chronic lung disease | 805,923 (17%) | 758,498 (16%) | 47,425 (42%) | 717,743 (16%) | 88,180 (35%) |
| Coronary artery disease | 358,693 (7.4%) | 321,889 (6.8%) | 36,804 (32%) | 300,058 (6.6%) | 58,635 (23%) |
| Dementia before | 91,768 (1.9%) | 74,969 (1.6%) | 16,799 (15%) | 72,777 (1.6%) | 18,991 (7.5%) |
| Depression | 775,932 (16%) | 743,839 (16%) | 32,093 (28%) | 712,396 (16%) | 63,536 (25%) |
| Diabetes | 694,589 (14%) | 649,536 (14%) | 45,053 (40%) | 603,168 (13%) | 91,421 (36%) |
| Heart failure | 276,943 (5.7%) | 237,156 (5.0%) | 39,787 (35%) | 217,268 (4.8%) | 59,675 (23%) |
| HIV infection | 26,330 (0.5%) | 25,301 (0.5%) | 1,029 (0.9%) | 23,935 (0.5%) | 2,395 (0.9%) |
| Hypertension | 1,467,196 (30%) | 1,384,800 (29%) | 82,396 (73%) | 1,307,118 (29%) | 160,078 (63%) |
| Kidney disease | 373,494 (7.7%) | 330,800 (7.0%) | 42,694 (38%) | 304,813 (6.7%) | 68,681 (27%) |
| Liver disease | 275,187 (5.7%) | 257,054 (5.5%) | 18,133 (16%) | 242,741 (5.3%) | 32,446 (13%) |
| Myocardial infraction | 148,666 (3.1%) | 128,986 (2.7%) | 19,680 (17%) | 116,374 (2.5%) | 32,292 (13%) |
| Peripheral vascular disease | 170,530 (3.5%) | 150,385 (3.2%) | 20,145 (18%) | 140,038 (3.1%) | 30,492 (12%) |
| Rheumatologic disease | 271,327 (5.6%) | 253,266 (5.4%) | 18,061 (16%) | 238,991 (5.2%) | 32,336 (13%) |

Among patients with an RD diagnosis, 16% experienced a life-threatening COVID-19 outcome while 6.1% did not (Pearson’s χ2 test p < 0.001). Similarly, 13% of RD patients were hospitalized while 6.0% of RD patients were not (Pearson’s χ2 test p < 0.001) (all differences were statistically significant with Pearson’s χ2 test < 0.001 *p* < 0.001) (**Table 1**). The most prevalent RD classes, irrespective of association with outcomes, were rare neurologic diseases (n=78,460), rare systemic or rheumatologic diseases (n=47,172), and rare hematologic diseases (n=36,462) (**Figure 2**).

**Figure 2:**
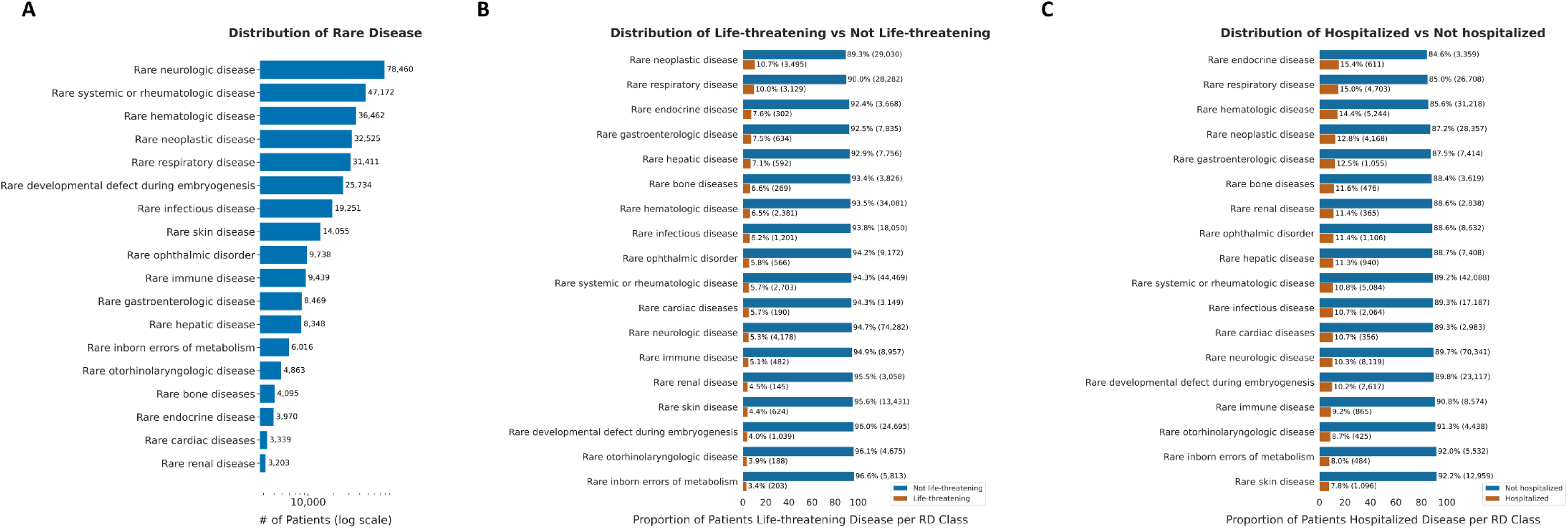
Description of patients with preexisting RD classes and associated life-threatening disease/ hospitalization in the N3C COVID-19 cohort. A) Number of patients per RD class identified within the N3C COVID-19 cohort. Proportion of RD patients, by RD class, in the N3C stratified by COVID-19 related life-threatening disease (B) and hospitalization (C).

The percent RD patients affected by life-threatening and hospitalization outcomes were further evaluated by Orphanet linearization class. The highest proportions of life-threatening outcomes were observed in patients with rare neoplastic diseases (10.7%; N=3,495) and rare respiratory diseases (10.0%; N=3,129). Hospitalization was most frequent among patients with rare respiratory diseases (15.0%, N=4,703) and rare hematologic diseases (14.4%; N=5,244) (**Figure 2**).

### Specific RD classes are associated with COVID-19 outcomes

In univariate logistic regression models, all 18 RD classes were significantly associated with increased odds ratios of life-threatening COVID-19, with the strongest associations observed for rare neoplastic (OR 5.12; 95% CI 4.95-5.31; *p* < 0.001), respiratory (OR 4.69; 95% CI 4.52-4.87; *p* < 0.001), and endocrine diseases (OR 3.42; 95% CI 3.04-3.85; *p* < 0.001). When examining COVID-19-related hospitalizations, the top three disease categories with the most robust associations were rare endocrine diseases (OR 3.27; 95% CI 3.0-3.56; *p* < 0.001), rare respiratory diseases (OR 3.2; 95% CI 3.1-3.3; *p* < 0.001), and rare hematologic diseases (OR 3.06; 95% CI 2.97-3.15; *p* < 0.001). (**Table S1**).

When adjusting for demographic and comorbidities variables, multivariate logistic regression models (**Table 2**, **Figure 3**) revealed that rare otorhinolaryngologic diseases were associated with the highest odds ratios of life-threatening COVID-19 outcomes (OR 4.51; 95% CI 3.81-5.33; *p* < 0.001), followed by rare developmental defects during embryogenesis (OR 1.84; 95% CI 1.72-1.98; *p* < 0·001), rare cardiac diseases (OR 1.79; 95% CI 1.51-2.11; *p* < 0·001), and rare respiratory diseases (OR 1.78; 95% CI 1.71-1.86; *p* < 0·001). All other RD classes were significantly associated with increased odds ratios of life-threatening illness (ORs range from 1.08 to 1.69), except for rare skin diseases (OR 0.98; 95% CI 0.90-1.07; *p* = 0.644), rare inborn errors of metabolism, and rare renal disease.

**Figure 3:**
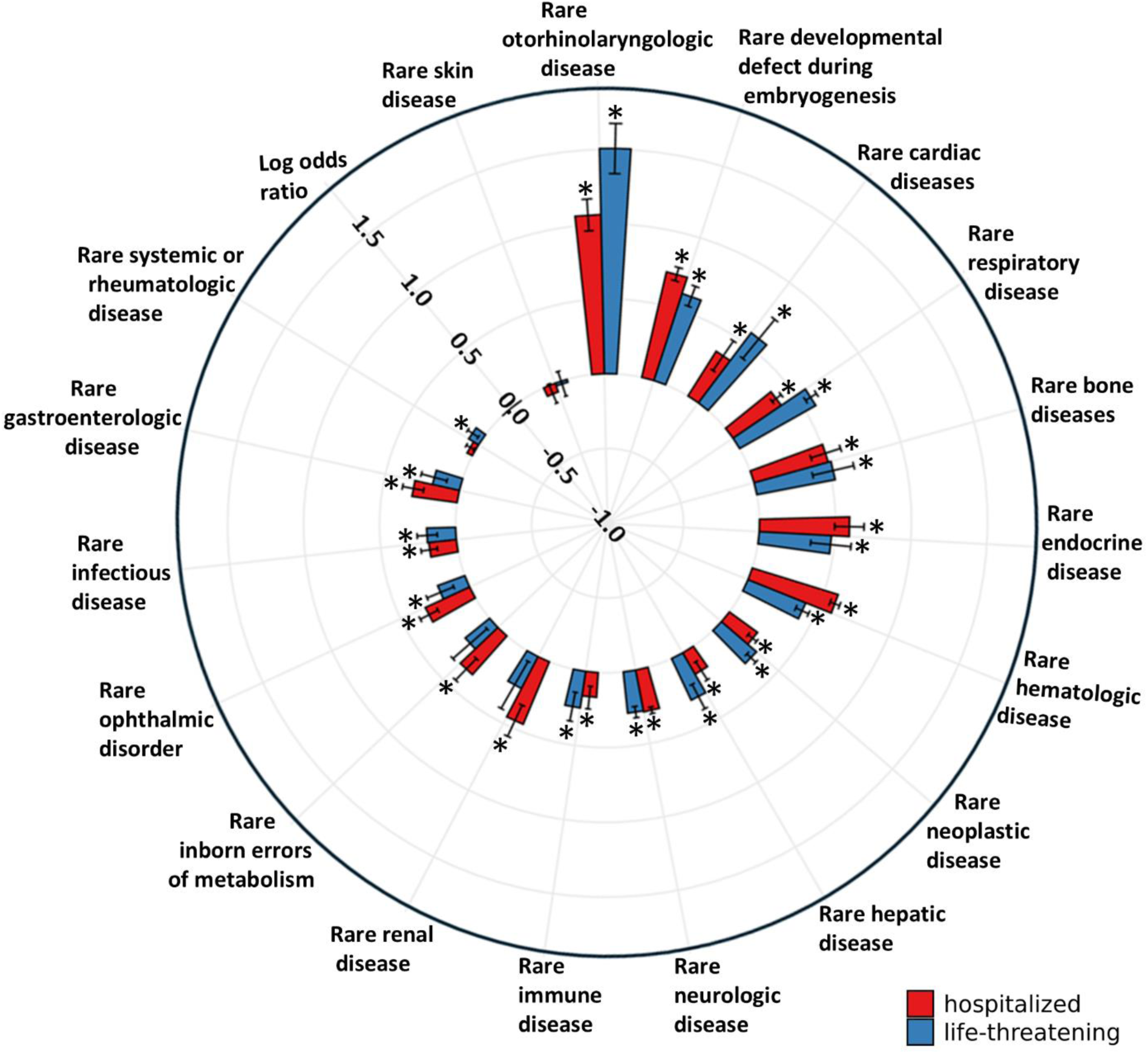
Multivariate weighted logistic regression models of each rare disease class association with COVID-19 life-threatening disease and hospitalization. Odds ratios for patients with vs. without preexisting RD for each RD class are depicted. Multivariate weighted logistic regression models were adjusted for demographics and comorbidities. Outcomes evaluated are hospitalization (red) and life-threatening disease (blue). * Denotes statistical significance (p-value < 0.0028).

**Table 2.** Multivariate logistic regression models of severity outcomes adjusted for demographics and preexisting comorbidities for each RD linearization class.

|  | Life-threatening |  |  | Hospitalized |  |
| --- | --- | --- | --- | --- | --- |
|  | OR (95% CI) | P value |  | OR (95% CI) | P value |
| Rare otorhinolaryngologic disease | **4.51 (3.81-5.33)** | **<0.001** | Rare otorhinolaryngologic disease | **2.90 (2.61-3.23)** | **<0.001** |
| Rare developmental defect during embryogenesis | **1.84 (1.72-1.98)** | **<0.001** | Rare developmental defect during embryogenesis | **2.06 (1.97-2.16)** | **<0.001** |
| Rare cardiac diseases | **1.79 (1.51-2.11)** | **<0.001** | Rare hematologic disease | **1.81 (1.75-1.87)** | **<0.001** |
| Rare respiratory disease | **1.78 (1.71-1.86)** | **<0.001** | Rare endocrine disease | **1.81 (1.64-1.99)** | **<0.001** |
| Rare bone diseases | **1.69 (1.47-1.94)** | **<0.001** | Rare bone diseases | **1.66 (1.50-1.84)** | **<0.001** |
| Rare endocrine disease | **1.61 (1.41-1.84)** | **<0.001** | Rare renal disease | **1.57 (1.40-1.76)** | **<0.001** |
| Rare hematologic disease | **1.51 (1.44-1.58)** | **<0.001** | Rare respiratory disease | **1.47 (1.42-1.53)** | **<0.001** |
| Rare neoplastic disease | **1.36 (1.31-1.42)** | **<0.001** | Rare cardiac diseases | **1.42 (1.26-1.60)** | **<0.001** |
| Rare hepatic disease | **1.36 (1.24-1.49)** | **<0.001** | Rare inborn errors of metabolism | **1.41 (1.28-1.56)** | **<0.001** |
| Rare neurologic disease | **1.32 (1.27-1.37)** | **<0.001** | Rare ophthalmic disorder | **1.38 (1.29-1.47)** | **<0.001** |
| Rare immune disease | **1.28 (1.16-1.41)** | **<0.001** | Rare gastroenterologic disease | **1.35 (1.26-1.45)** | **<0.001** |
| Rare renal disease | 1.27 (1.06-1.52) | 0.0092 | Rare neurologic disease | **1.33 (1.30-1.36)** | **<0.001** |
| Rare inborn errors of metabolism | 1.23 (1.06-1.43) | 0.0079 | Rare neoplastic disease | **1.26 (1.21-1.30)** | **<0.001** |
| Rare infectious disease | **1.21 (1.13-1.29)** | **<0.001** | Rare infectious disease | **1.20 (1.14-1.27)** | **<0.001** |
| Rare ophthalmic disorder | **1.21 (1.10-1.33)** | **<0.001** | Rare immune disease | **1.18 (1.10-1.28)** | **<0.001** |
| Rare gastroenterologic disease | **1.20 (1.10-1.31)** | **<0.001** | Rare hepatic disease | **1.18 (1.10-1.27)** | **<0.001** |
| Rare systemic or rheumatologic disease | **1.08 (1.03-1.12)** | **<0.001** | Rare systemic or rheumatologic disease | 1.04 (1.01-1.07) | 0.0135 |
| Rare skin disease | 0.98 (0.90-1.07) | 0.6440 | Rare skin disease | 0.94 (0.88-1.01) | 0.0704 |

Similar to life-threatening disease, adjusted multivariate regression models evaluating COVID-19 related hospitalization show that rare otorhinolaryngologic diseases (OR 2.90; 95% CI 2.61-3.23), rare developmental defect during embryogenesis (OR 2.06; 95% CI 1.97-2.16), and rare hematologic diseases (OR 1.81; 95% CI 1.75-1.87) are the top three RD classes showing the strongest positive association with increased risk (p < 0.001). All other classes were also associated with elevated odds ratios except rare skin diseases and rare systemic or rheumatologic diseases (**Table 2**, **Figure 3**).

### Vaccination and antiviral treatment exposures are protective in RD and non-RD populations

To assess the impact of vaccination and antiviral treatment, we performed a sub-cohort analysis using data from sites with reliable treatment documentation from December 23^rd^, 2021, to January 4^th^, 2024. Among 799,662 vaccinated and/or treated patients, 66,021 (8.3%) had a RD. Compared to non-RD, RD patients were more likely to receive both antivirals and vaccination compared to non-RD patients (19% vs. 16%, respectively, Pearson’s χ2 test < 0.001) and had higher rates of severe outcomes, including hospitalization (9.5% in RD patients vs. 4.5% in non-RD patients, Pearson’s χ2 test < 0.001) and long COVID (11% in RD patients vs. 5.4% in non-RD patients, Pearson’s χ2 test < 0.001). RD patients were also older and had a greater burden of comorbidities such as cancer, chronic lung disease, and hypertension compared to non-RD patients (all differences were statistically significant with Pearson’s χ2 test < 0.001 *p* < 0.001) (**Table S2**).

Among patients with preexisting RDs, COVID-19 vaccination alone was associated with a reduction in the odds of developing life-threatening disease compared to unvaccinated individuals (OR = 0.71; 95% CI: 0.66-0.77; *p* < 0.001) **(Table 3)**. Antiviral treatment alone was also protective (OR = 0.33; 95% CI: 0.26-0.42; *p* < 0.001), and the combination of vaccination and antiviral therapy conferred the strongest protection (OR = 0.24; 95% CI: 0.20-0.27; *p* < 0.001). Similarly, in patients without preexisting RDs, vaccination alone was associated with a reduction in the odds of life-threatening outcomes compared to no vaccination (OR = 0.64; 95% CI: 0.61-0.66; *p* < 0.001), antiviral treatment alone also conferred protection (OR = 0.34; 95% CI: 0.30-0.38; *p* < 0.001), and a combined vaccination and antiviral therapy showed the strongest protection effect (OR = 0.23; 95% CI: 0.21-0.25; *p* < 0.001) (**Table 3**). All models were adjusted for demographics and comorbidities. The protective effects of vaccination and antiviral treatment were thus consistent across patients with and without preexisting RD diagnosis, underscoring their benefit in medically vulnerable individuals.

**Table 3:** Impact of vaccination and antiviral treatment exposures on life-threatening disease in patients with and without preexising RDs (multivariate logistic regression models are adjusted for demographics and comorbidities).

|  | Patients with Preexisting Rare Disease |  |  |  |  |  | Patients without Preexisting Rare Disease |  |  |  |  |  |
| --- | --- | --- | --- | --- | --- | --- | --- | --- | --- | --- | --- | --- |
|  | Vaccination vs Control |  | Antiviral treatment vs Control |  | Vaccination + antiviral treatment vs Control |  | Vaccination vs Control |  | Antiviral treatment vs Control |  | Vaccination + antiviral treatment vs Control |  |
|  | Life-threatening condition<br>Yes: 2,942<br>No: 46,793 |  | Life-threatening condition<br>Yes: 1,439<br>No: 24,707 |  | Life-threatening condition<br>Yes: 3,269<br>No: 62,752 |  | Life-threatening condition<br>Yes: 12,068<br>No: 566,109 |  | Life-threatening condition<br>Yes: 6,568<br>No: 315,387 |  | Life-threatening condition<br>Yes: 13,443<br>No: 720,198 |  |
|  | OR (95% CI) | P value | OR (95% CI) | P value | OR (95% CI) | P value | OR (95% CI) | P value | OR (95% CI) | P value | OR (95% CI) | P value |
| **COVID-19 prevention/intervention** |  |  |  |  |  |  |  |  |  |  |  |  |
| Control | 1 [Reference] |  | 1 [Reference] |  | 1 [Reference] |  | 1 [Reference] |  | 1 [Reference] |  | 1 [Reference] |  |
| Vaccination status | **0.71 (0.66 - 0.77)** | **< 0.001** |  |  |  |  | **0.64 (0.61 - 0.66)** | **< 0.001** |  |  |  |  |
| Antiviral treatment |  |  | **0.33 (0.26 - 0.42)** | **< 0.001** |  |  |  |  | **0.34 (0.3 - 0.38)** | **< 0.001** |  |  |
| Vaccination & Antiviral |  |  |  |  | **0.24 (0.2 - 0.27)** | **< 0.001** |  |  |  |  | **0.23 (0.21 - 0.25)** | **< 0.001** |
| **Demographic** |  |  |  |  |  |  |  |  |  |  |  |  |
| **Age** |  |  |  |  |  |  |  |  |  |  |  |  |
| Age (20-40) | 1 [Reference] |  | 1 [Reference] |  | 1 [Reference] |  | 1 [Reference] |  | 1 [Reference] |  | 1 [Reference] |  |
| Age (1-20) | 0.67 (0.48 - 0.95) | 0.024 | 0.87 (0.62 - 1.24) | 0.444 | 0.92 (0.65 - 1.31) | 0.661 | 0.3 (0.24 - 0.38) | < 0.001 | 0.29 (0.23 - 0.37) | < 0.001 | 0.31 (0.24 - 0.4) | < 0.001 |
| Age (40-65) | 1.91 (1.59 - 2.29) | < 0.001 | 2.28 (1.81 - 2.86) | < 0.001 | 2.3 (1.82 - 2.9) | < 0.001 | 2.99 (2.71 - 3.3) | < 0.001 | 2.75 (2.45 - 3.08) | < 0.001 | 2.73 (2.43 - 3.07) | < 0.001 |
| Age (>65) | 3.05 (2.53 - 3.68) | < 0.001 | 3.46 (2.73 - 4.39) | < 0.001 | 3.51 (2.76 - 4.48) | < 0.001 | 6.35 (5.72 - 7.03) | < 0.001 | 5.59 (4.94 - 6.33) | < 0.001 | 5.61 (4.95 - 6.35) | < 0.001 |
| **BMI** |  |  |  |  |  |  |  |  |  |  |  |  |
| Normal | 1 [Reference] |  | 1 [Reference] |  | 1 [Reference] |  | 1 [Reference] |  | 1 [Reference] |  | 1 [Reference] |  |
| Obese | 0.56 (0.5 - 0.64) | < 0.001 | 0.55 (0.46 - 0.65) | < 0.001 | 0.58 (0.49 - 0.68) | < 0.001 | 0.56 (0.52 - 0.59) | < 0.001 | 0.56 (0.52 - 0.61) | < 0.001 | 0.56 (0.52 - 0.61) | < 0.001 |
| Over weight | 0.67 (0.59 - 0.76) | < 0.001 | 0.69 (0.58 - 0.83) | < 0.001 | 0.71 (0.6 - 0.85) | < 0.001 | 0.67 (0.63 - 0.72) | < 0.001 | 0.69 (0.63 - 0.75) | < 0.001 | 0.68 (0.63 - 0.74) | < 0.001 |
| Under weight | 1.2 (0.83 - 1.73) | 0.335 | 1 (0.68 - 1.47) | 0.994 | 0.99 (0.67 - 1.46) | 0.953 | 1.47 (1.22 - 1.77) | < 0.001 | 1.21 (0.98 - 1.49) | 0.079 | 1.22 (0.98 - 1.51) | 0.079 |
| **Sex** |  |  |  |  |  |  |  |  |  |  |  |  |
| Female | 1 [Reference] |  | 1 [Reference] |  | 1 [Reference] |  | 1 [Reference] |  | 1 [Reference] |  | 1 [Reference] |  |
| Male | 1.52 (1.39 - 1.66) | < 0.001 | 1.48 (1.32 - 1.67) | < 0.001 | 1.49 (1.32 - 1.67) | < 0.001 | 1.39 (1.34 - 1.45) | < 0.001 | 1.43 (1.35 - 1.51) | < 0.001 | 1.4 (1.33 - 1.48) | < 0.001 |
| **Race** |  |  |  |  |  |  |  |  |  |  |  |  |
| White | 1 [Reference] |  | 1 [Reference] |  | 1 [Reference] |  | 1 [Reference] |  | 1 [Reference] |  | 1 [Reference] |  |
| Asian | 0.82 (0.65 - 1.04) | 0.102 | 0.86 (0.63 - 1.17) | 0.338 | 0.77 (0.56 - 1.06) | 0.106 | 0.75 (0.68 - 0.84) | < 0.001 | 0.73 (0.64 - 0.84) | < 0.001 | 0.77 (0.67 - 0.88) | < 0.001 |
| Black or African American | 1.21 (1.07 - 1.36) | 0.002 | 1.31 (1.11 - 1.55) | 0.001 | 1.24 (1.05 - 1.46) | 0.011 | 1.16 (1.09 - 1.23) | < 0.001 | 1.22 (1.12 - 1.32) | < 0.001 | 1.18 (1.09 - 1.29) | < 0.001 |
| Missing/Unknown/Other | 1.07 (0.9 - 1.28) | 0.453 | 1.03 (0.82 - 1.3) | 0.779 | 1.05 (0.83 - 1.31) | 0.690 | 1.07 (0.99 - 1.16) | 0.098 | 1.07 (0.97 - 1.19) | 0.177 | 1.11 (1 - 1.23) | 0.042 |
| **Ethnicity** |  |  |  |  |  |  |  |  |  |  |  |  |
| Not hispanic or Latino | 1 [Reference] |  | 1 [Reference] |  | 1 [Reference] |  | 1 [Reference] |  | 1 [Reference] |  | 1 [Reference] |  |
| Hispanic or Latino | 1.01 (0.81 - 1.25) | 0.952 | 1.21 (0.93 - 1.58) | 0.151 | 1.3 (1.01 - 1.69) | 0.045 | 0.93 (0.85 - 1.03) | 0.153 | 1.01 (0.9 - 1.14) | 0.836 | 1.01 (0.89 - 1.13) | 0.916 |
| Missing/Unknown | 0.72 (0.51 - 1.02) | 0.065 | 0.6 (0.37 - 0.99) | 0.044 | 0.58 (0.35 - 0.95) | 0.029 | 0.66 (0.56 - 0.77) | < 0.001 | 0.7 (0.57 - 0.85) | < 0.001 | 0.66 (0.54 - 0.81) | < 0.001 |
| **Smoking status** |  |  |  |  |  |  |  |  |  |  |  |  |
| Non-smoker | 1 [Reference] |  | 1 [Reference] |  | 1 [Reference] |  | 1 [Reference] |  | 1 [Reference] |  | 1 [Reference] |  |
| Current or former smoker | 0.95 (0.85 - 1.06) | 0.370 | 0.78 (0.66 - 0.91) | 0.002 | 0.8 (0.68 - 0.93) | 0.004 | 1.05 (1 - 1.11) | 0.050 | 0.97 (0.91 - 1.05) | 0.489 | 0.96 (0.9 - 1.04) | 0.306 |
| **Comorbidities** |  |  |  |  |  |  |  |  |  |  |  |  |
| Cancer | 2.11 (1.93 - 2.3) | < 0.001 | 2.26 (2 - 2.56) | < 0.001 | 2.32 (2.06 - 2.61) | < 0.001 | 2.57 (2.45 - 2.69) | < 0.001 | 2.83 (2.65 - 3.01) | < 0.001 | 2.8 (2.63 - 2.98) | < 0.001 |
| Cardiomyopathies | 1.02 (0.9 - 1.15) | 0.788 | 0.94 (0.78 - 1.13) | 0.501 | 0.94 (0.79 - 1.12) | 0.509 | 1.01 (0.94 - 1.09) | 0.745 | 0.99 (0.89 - 1.09) | 0.767 | 0.98 (0.89 - 1.08) | 0.662 |
| Cerebro vascular disease | 1.13 (1.02 - 1.26) | 0.018 | 1.17 (1 - 1.36) | 0.048 | 1.14 (0.98 - 1.32) | 0.087 | 1.17 (1.1 - 1.24) | < 0.001 | 1.17 (1.08 - 1.27) | < 0.001 | 1.16 (1.07 - 1.25) | < 0.001 |
| Chronic lung disease | 1.4 (1.28 - 1.54) | 0.000 | 1.34 (1.18 - 1.52) | < 0.001 | 1.34 (1.18 - 1.51) | < 0.001 | 1.62 (1.55 - 1.69) | < 0.001 | 1.59 (1.5 - 1.69) | < 0.001 | 1.61 (1.51 - 1.7) | < 0.001 |
| Coronary artery disease | 1.01 (0.9 - 1.13) | 0.875 | 0.95 (0.81 - 1.11) | 0.524 | 1.01 (0.87 - 1.18) | 0.851 | 1.03 (0.97 - 1.09) | 0.347 | 0.99 (0.92 - 1.07) | 0.808 | 0.99 (0.92 - 1.07) | 0.806 |
| Dementia before | 2.18 (1.91 - 2.49) | < 0.001 | 2.21 (1.83 - 2.67) | < 0.001 | 2.22 (1.84 - 2.66) | < 0.001 | 3.09 (2.9 - 3.29) | < 0.001 | 2.84 (2.6 - 3.1) | < 0.001 | 3.06 (2.81 - 3.34) | < 0.001 |
| Depression | 1.01 (0.92 - 1.11) | 0.798 | 1.04 (0.92 - 1.19) | 0.525 | 1.01 (0.89 - 1.15) | 0.844 | 1.04 (0.99 - 1.09) | 0.111 | 1.07 (1 - 1.15) | 0.036 | 1.06 (1 - 1.14) | 0.068 |
| Diabetes | 1.07 (0.97 - 1.17) | 0.199 | 1.05 (0.92 - 1.21) | 0.468 | 1.14 (0.99 - 1.3) | 0.064 | 1.16 (1.11 - 1.22) | < 0.001 | 1.15 (1.07 - 1.23) | < 0.001 | 1.15 (1.08 - 1.23) | < 0.001 |
| Heart failure | 2.09 (1.87 - 2.34) | < 0.001 | 1.95 (1.67 - 2.29) | < 0.001 | 2.02 (1.73 - 2.35) | < 0.001 | 2.2 (2.08 - 2.34) | < 0.001 | 2.08 (1.92 - 2.26) | < 0.001 | 2.15 (1.98 - 2.33) | < 0.001 |
| HIV infection | 0.96 (0.7 - 1.31) | 0.783 | 1.09 (0.73 - 1.63) | 0.686 | 1.12 (0.75 - 1.67) | 0.574 | 0.98 (0.77 - 1.24) | 0.839 | 0.93 (0.67 - 1.29) | 0.659 | 0.97 (0.7 - 1.35) | 0.863 |
| Hypertension | 1.2 (1.06 - 1.35) | 0.004 | 1.31 (1.11 - 1.55) | 0.001 | 1.27 (1.08 - 1.49) | 0.004 | 1.39 (1.31 - 1.47) | < 0.001 | 1.36 (1.26 - 1.47) | < 0.001 | 1.31 (1.21 - 1.41) | < 0.001 |
| Kidney disease | 1.73 (1.57 - 1.9) | < 0.001 | 1.75 (1.53 - 1.99) | < 0.001 | 1.64 (1.44 - 1.86) | < 0.001 | 1.83 (1.75 - 1.93) | < 0.001 | 1.8 (1.68 - 1.93) | < 0.001 | 1.82 (1.7 - 1.94) | < 0.001 |
| Liver disease | 1.42 (1.29 - 1.57) | < 0.001 | 1.41 (1.23 - 1.62) | < 0.001 | 1.39 (1.22 - 1.59) | < 0.001 | 1.43 (1.34 - 1.52) | < 0.001 | 1.44 (1.33 - 1.56) | < 0.001 | 1.39 (1.28 - 1.5) | < 0.001 |
| Myocardial infraction | 0.99 (0.88 - 1.12) | 0.866 | 0.88 (0.74 - 1.05) | 0.148 | 0.91 (0.77 - 1.08) | 0.298 | 1.2 (1.13 - 1.28) | < 0.001 | 1.22 (1.12 - 1.33) | < 0.001 | 1.25 (1.15 - 1.36) | < 0.001 |
| Peripheral vascular disease | 1.18 (1.06 - 1.32) | 0.003 | 1.1 (0.94 - 1.3) | 0.238 | 1.13 (0.97 - 1.33) | 0.119 | 1.19 (1.12 - 1.26) | < 0.001 | 1.2 (1.1 - 1.3) | < 0.001 | 1.18 (1.09 - 1.29) | < 0.001 |
| Rheumatologic disease | 0.93 (0.85 - 1.02) | 0.143 | 0.86 (0.75 - 0.99) | 0.033 | 0.9 (0.79 - 1.03) | 0.135 | 1 (0.95 - 1.06) | 0.952 | 0.99 (0.91 - 1.08) | 0.839 | 1.01 (0.93 - 1.09) | 0.860 |

Notably, the results for hospitalization mirrored those observed for life-threatening outcomes. Vaccination (OR 0.62; 95% CI: 95% CI: 0.59-0.66; p < 0.001), antiviral treatment (OR 0.31; 95% CI: 95% CI: 0.26-0.37; *p* < 0.001), and the combined use of both (OR 0.11; 95% CI: 95% CI: 0.10-0.13; *p* < 0.001) showed substantial and statistically significant protective effects of hospitalization. Similar protection was noted in patients without RDs (**Table S3**).

### Vaccination and antiviral treatment exposure show no impact on long COVID outcome in RD patient population

We evaluated whether vaccination or antiviral treatment (Paxlovid and Molnupiravir) are associated with long COVID in RD and non-RD patients (**Table 4**). In patients with preexisting RD, vaccination alone (OR: 1.09; 95% CI: 1.02-1.16; *p* = 0.006) and antiviral treatment alone (OR: 1.01; 95% CI: 0.89-1.14; *p* = 0.93) was not protective of long COVID. However, the combination of vaccination and antivirals did show a protective effect (OR: 0.88; 95% CI: 0.81-0.96; *p* = 0.002). Similarly, patients without RDs showed a protective effect against long COVID for patients that underwent vaccination and antiviral therapy both (OR: 0.84; 95% CI: 0.81-0.87; *p* < 0.001). Contrary to non-RD patients, the protective effect was not statistically significant for vaccination (OR: 1.09; 95% CI: 1.02-1.16; *p* = 0.006) alone or antiviral treatment (OR: 1.01; 95% CI: 0.89-1.14; *p =* 0.93) alone for RD patients.

**Table 4:**
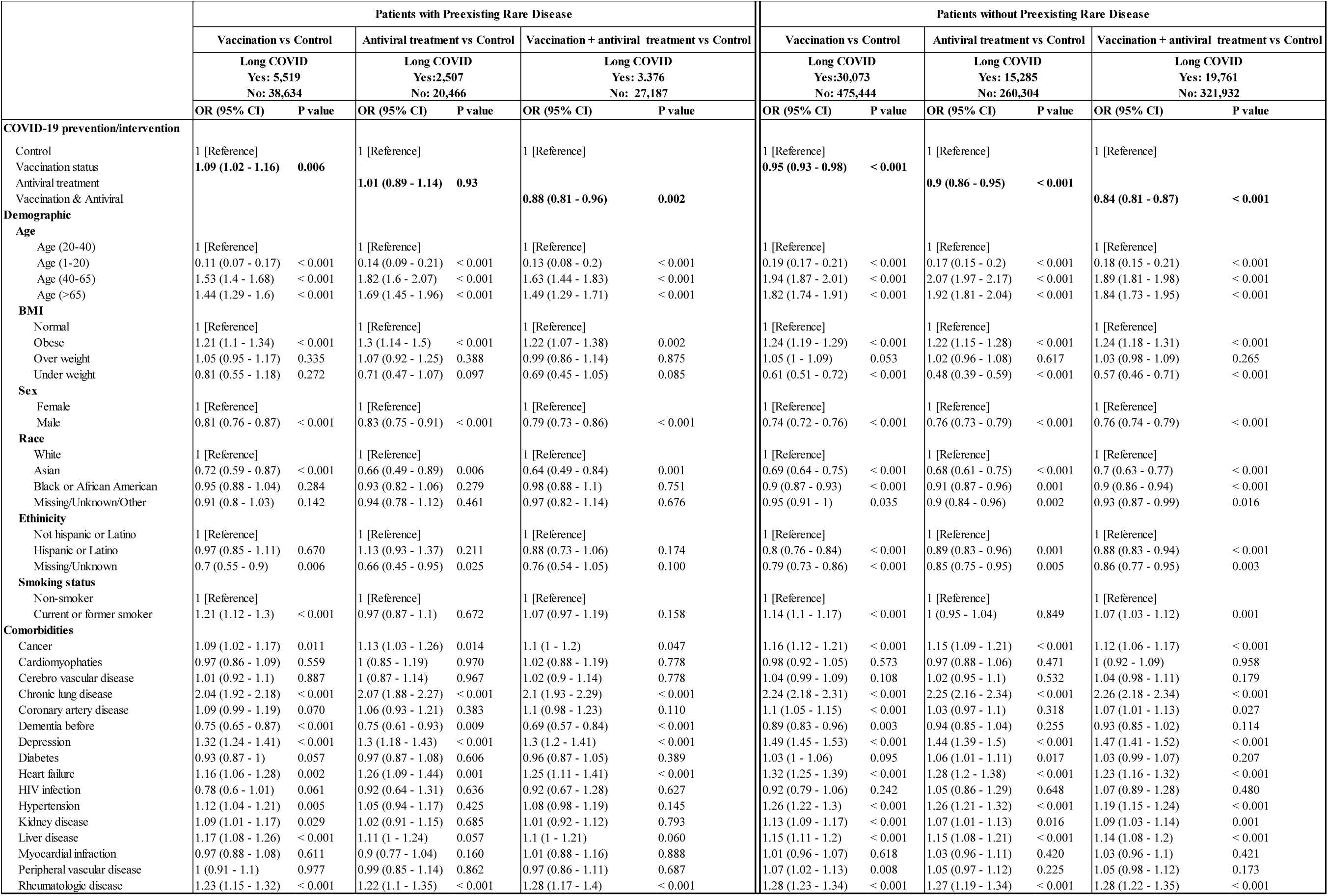
Impact of vaccination and antiviral treatment exposures on long-COVID in patients with and without preexising RDs (multivariate logistic regression models are adjusted for demographics and comorbidities).

### Protective effect of vaccination on COVID reinfection

Vaccination was associated with a significant reduction in the risk of COVID-19 reinfection in individuals with and without preexisting RDs (**Table 5**). Among 64,128 RD patients, vaccinated individuals had had lower odds of reinfection compared to unvaccinated controls (OR 0.81; 95% CI 0.75-0.86; *p* < 0.001). Similarly, in the larger non-RD population of 804,853 individuals, vaccination conferred an even greater protective effect (OR 0.75; 95% CI 0.73-0.76; *p* < 0.001). These findings demonstrate that while vaccination provides meaningful protection against reinfection across both groups, the effect is somewhat more pronounced in those without RD. We also note that age was inversely associated with reinfection risk in both RD and non-RD patients, with older individuals experiencing lower risk compared to those aged 20-40 years. Overall, these results support the continued prioritization of vaccination efforts to reduce COVID-19 reinfections, especially among vulnerable RD patients who face elevated risks from the virus.

**Table 5:** Impact of vaccination on COVID-19 reinfection in patients with and without preexisting RDs (multivariate logistic regression models adjusted for demographics and comorbidities).

|  | Patients with Preexisting Rare Disease |  | Patients without Preexisting Rare Disease |  |
| --- | --- | --- | --- | --- |
|  | COVID Reinfection |  | COVID Reinfection |  |
|  | Yes: 4,066<br>No: 60,062 |  | Yes: 40,466<br>No: 764,387 |  |
|  | OR (95% CI) | P value | OR (95% CI) | P value |
| **COVID-19 prevention/intervention** |  |  |  |  |
| Control | 1 [Reference] |  | 1 [Reference] |  |
| Vaccination status | **0.81 (0.75 - 0.86)** | **< 0.001** | **0.75 (0.73 - 0.76)** | **< 0.001** |
| **Demographic** |  |  |  |  |
| **Age** |  |  |  |  |
| Age (20-40) | 1 [Reference] |  | 1 [Reference] |  |
| Age (1-20) | 0.81 (0.7 - 0.95) | 0.008 | 0.86 (0.83 - 0.89) | < 0.001 |
| Age (40-65) | 0.82 (0.75 - 0.89) | < 0.001 | 0.81 (0.79 - 0.83) | < 0.001 |
| Age (>65) | 0.67 (0.6 - 0.75) | < 0.001 | 0.65 (0.62 - 0.67) | < 0.001 |
| **BMI** |  |  |  |  |
| Normal | 1 [Reference] |  | 1 [Reference] |  |
| Obese | 1.04 (0.94 - 1.15) | 0.427 | 1.23 (1.19 - 1.27) | < 0.001 |
| Over weight | 0.97 (0.86 - 1.08) | 0.541 | 1.09 (1.05 - 1.13) | < 0.001 |
| Under weight | 0.88 (0.7 - 1.09) | 0.237 | 0.89 (0.84 - 0.95) | < 0.001 |
| **Sex** |  |  |  |  |
| Female | 1 [Reference] |  | 1 [Reference] |  |
| Male | 0.91 (0.85 - 0.98) | 0.012 | 0.86 (0.84 - 0.88) | < 0.001 |
| **Race** |  |  |  |  |
| White | 1 [Reference] |  | 1 [Reference] |  |
| Asian | 0.93 (0.74 - 1.17) | 0.530 | 0.91 (0.85 - 0.96) | 0.002 |
| Black or African American | 1.11 (1.01 - 1.21) | 0.026 | 1.16 (1.13 - 1.2) | < 0.001 |
| Missing/Unknown/Other | 1.01 (0.89 - 1.15) | 0.865 | 1.06 (1.02 - 1.1) | 0.001 |
| **Ethnicity** |  |  |  |  |
| Not hispanic or Latino | 1 [Reference] |  | 1 [Reference] |  |
| Hispanic or Latino | 0.99 (0.86 - 1.14) | 0.895 | 1.03 (1 - 1.07) | 0.054 |
| Missing/Unknown | 0.64 (0.49 - 0.85) | 0.002 | 0.69 (0.65 - 0.74) | < 0.001 |
| **Smoking status** |  |  |  |  |
| Non-smoker | 1 [Reference] |  | 1 [Reference] |  |
| Current or former smoker | 0.85 (0.77 - 0.93) | < 0.001 | 0.84 (0.82 - 0.87) | < 0.001 |
| **Comorbidities** |  |  |  |  |
| Cancer | 1.3 (1.2 - 1.4) | < 0.001 | 1.05 (1.01 - 1.09) | 0.016 |
| Cardiomyopathies | 0.88 (0.76 - 1.01) | 0.071 | 1.02 (0.95 - 1.1) | 0.593 |
| Cerebro vascular disease | 1 (0.9 - 1.11) | 0.981 | 1.07 (1.01 - 1.13) | 0.019 |
| Chronic lung disease | 1.18 (1.1 - 1.27) | < 0.001 | 1.16 (1.13 - 1.19) | < 0.001 |
| Coronary artery disease | 1.13 (1.01 - 1.25) | 0.030 | 1.09 (1.04 - 1.15) | < 0.001 |
| Dementia before | 1.25 (1.07 - 1.45) | 0.005 | 1.12 (1.03 - 1.2) | 0.005 |
| Depression | 1.04 (0.97 - 1.12) | 0.262 | 1.07 (1.05 - 1.1) | < 0.001 |
| Diabetes | 1.02 (0.94 - 1.11) | 0.669 | 1.01 (0.98 - 1.05) | 0.434 |
| Heart failure | 1.13 (1.01 - 1.27) | 0.035 | 1.11 (1.04 - 1.17) | 0.001 |
| HIV infection | 0.98 (0.76 - 1.28) | 0.891 | 0.85 (0.74 - 0.98) | 0.024 |
| Hypertension | 1.06 (0.97 - 1.15) | 0.215 | 1.02 (0.99 - 1.05) | 0.207 |
| Kidney disease | 1.1 (1 - 1.2) | 0.040 | 1.12 (1.07 - 1.16) | < 0.001 |
| Liver disease | 1.19 (1.09 - 1.3) | < 0.001 | 1.11 (1.07 - 1.16) | < 0.001 |
| Myocardial infraction | 1.04 (0.92 - 1.18) | 0.484 | 1.11 (1.04 - 1.18) | 0.001 |
| Peripheral vascular disease | 0.94 (0.83 - 1.06) | 0.312 | 1.09 (1.03 - 1.15) | 0.004 |
| Rheumatologic disease | 1.09 (1.01 - 1.19) | 0.036 | 1.11 (1.06 - 1.16) | < 0.001 |

## Discussion

This study leveraging a large-scale real-world dataset from over 21 million individuals in the N3C enclave provides the most comprehensive assessment of COVID-19 severity, reinfection, and long COVID outcomes among patients with RDs. Our findings underscore the disproportionate burden of severe COVID-19 outcomes in this medically vulnerable population and offer crucial insights into the protective role of vaccination and antiviral therapy.

Patients with RDs experienced markedly worse outcomes following SARS-CoV-2 infection compared to those without RDs. Life-threatening diseases and hospitalization occurred at more than twice the rate of the RD population. Notably, among the 18 RDs classes examined, nearly all showed significantly elevated odds ratios of severe outcomes, with rare otorhinolaryngologic, developmental defect during embryogenesis, cardiac, respiratory diseases, hematologic, and endocrine conferring the highest risks. For patients with otorhinolaryngologic conditions, their association with worse outcomes could be explained in part by poor clearance of secretions or aspiration that might favor the development of pneumonia. According to the CDC, developmental, cardiac, and respiratory disorders are classified as disabilities that increase the risk of COVID-19 and some of these disorders include RDs (e.g., Cystic fibrosis, Gaucher disease, Fragile X syndrome etc.)^29^.

These findings echo existing literature highlighting the importance of underlying conditions in COVID-19 prognosis, including cardiopulmonary disease, disabilities, cancer, liver, kidney, and neurologic disease and immunologic impairments^30,31^. Our findings also expand this risk paradigm to previously understudied RD groups (e.g., patients with rare otorhinolaryngologic, bone, and endocrine disease), in which the underlying basis for elevated risk may be less clear and deserving of further study.

Importantly, these associations persisted even after controlling for common comorbidities and demographic factors that are known to increase the risk of adverse outcomes, suggesting that RDs contribute independently to vulnerability. This has significant implications for clinical risk stratification, particularly since many RDs lack specific public health guidelines for pandemic preparedness and response.

Encouragingly, we observed that both COVID-19 vaccination and antiviral treatment were highly effective in reducing the risk of severe illness in patients with RDs, mirroring trends seen in the general, non-RD population. The combined effect of both interventions was particularly notable, yielding an odds ratio of 0.24 for life-threatening outcomes and 0.11 for hospitalization when compared to no intervention. These findings support and extend prior work demonstrating the additive benefits of layered COVID-19 prevention strategies^32,33^ and reaffirm the importance of timely antiviral treatment in high-risk groups^34^.

However, the protective benefits of antiviral treatment and vaccination exposure was not uniform across all outcomes. While vaccination and/or antivirals substantially mitigated the risk of acute severity compared to no intervention, they offered no significant protection against long COVID in patients with RDs when administered alone. This contrasts with findings in the non-RD population, where vaccination alone or antiviral treatment alone reduced long COVID risk compared to no treatment. These differential effects raise important questions about the pathophysiology of post-acute sequelae in RD populations. It is possible that overlapping symptomatology, pre-existing organ dysfunction, or immune dysregulation in RD patients may obscure or complicate recovery trajectories, limiting the measurable impact of current interventions^35–37^.

Similarly, our findings on COVID-19 reinfection reveal that vaccination provides significant protection in both RD and non-RD populations, though the magnitude of protection is slightly attenuated in individuals with RDs (OR = 0.81 for RD patients and OR = 0.75 in non-RD patients). This suggests a nuanced protective profile, possibly due to differential immune responses in RD individuals, particularly those with immune or hematologic impairments. Antiviral treatments were not explicitly analyzed for their effect on reinfection because vaccination was widely implemented starting in late 2020 to early 2021, whereas antiviral therapies received approval only in late 2021, limiting the observation window to assess their impact on reinfection rates. Nonetheless, the demonstrated protective effect of vaccination underscores the importance of tailored preventive strategies to enhance protection in vulnerable RD populations.

This study has several strengths, including its unprecedented scale, disease-specific stratification using Orphanet classification, and robust confounder adjustment. However, some limitations must be acknowledged. Our reliance on EHR data may undercapture long COVID and reinfection events, particularly those not resulting in healthcare encounters. Furthermore, cases of long COVID were also identified in our analysis using a machine learning model that has not been specifically validated in a population of patients with RDs, in which chronic and recurrent symptoms might overlap with the clinical picture of long COVID. The heterogeneity within and between RD classes may also limit generalizability, and concept mapping limitations necessitated the exclusion of 12 RD classes. Future research should incorporate patient-reported outcomes, immunologic profiling, and mechanistic studies to better understand the differential impacts of COVID-19 in these populations and specific factors that may predispose to worse outcomes.

In conclusion, our findings highlight the heightened vulnerability of patients with RD to COVID-19 severity, long-COVID, and reinfection, while underscoring the effectiveness of combined vaccination and antiviral therapy to protect against long-COVID, and of vaccination to protect against reinfection. Tailored public health strategies, prioritization in treatment allocation, and dedicated clinical trials are essential to protect these often-overlooked populations in the ongoing pandemic and beyond.

## Data Availability

All data produced are available online at

https://covid.cd2h.org

## Contributors

E.A.M. and A.S.Y. conceptualized the original idea and designed the study. A.S.Y. and E.A.M. had full access to all the data in the study. A.S.Y. performed all data analyses. A.S.Y., D.K.S., and E.A.M. interpreted the results and take responsibility for the accuracy of the analyses and results. A.S.Y. and E.A.M drafted the manuscript. D.K.S. and E.Y.C. provided input on ophthalmic rare disease classification. E.S. and D.P. served as subject matter experts on rare disease classification. All authors reviewed and approved the final manuscript and agreed to be accountable for all aspects of the work.

## Data sharing

The N3C data transfer to NCATS is performed under a Johns Hopkins University Reliance Protocol # IRB00249128 or individual site agreements with NIH. The N3C Data Enclave is managed under the authority of the NIH; information can be found at https://ncats.nih.gov/n3c/resources. Enclave data is protected, to access N3C EHR data, researchers must register, ensure their institution has a Data Use Agreements (DUA) with NCATS, complete data security training, and submit a DUR for DAC approval. Enclave and data access instructions can be found at https://covid.cd2h.org/for-researchers.. The analytical code of this project is available at https://github.com/arjunyadaw/Rare-Disease-long-COVID-COVID-Reinfection-N3C-Enclave.git.

## N3C Acknowledgements

### National COVID Cohort Collaborative attribution

The analyses described in this [abstract/publication/report/presentation] were conducted with data or tools accessed through the NCATS N3C Data Enclave https://covid.cd2h.org and N3C Attribution & Publication Policy v 1.2-2020-08-25b supported by NCATS Contract No. 75N95023D00001, Axle Informatics Subcontract: NCATS-P00438-B, and [insert additional funding agencies or sources and reference numbers as declared by the contributors in their form response above]. This research was possible because of the patients whose information is included within the data and the organizations (https://ncats.nih.gov/n3c/resources/data-contribution/data-transfer-agreement-signatories) and scientists who have contributed to the on-going development of this community resource [https://doi.org/10.1093/jamia/ocaa196].

#### Disclaimer

The N3C Publication committee confirmed that this manuscript MSID: 2490.893 is in accordance with N3C data use and attribution policies; however, this content is solely the responsibility of the authors and does not necessarily represent the official views of the National Institutes of Health or the N3C program.

### Individual acknowledgments for core contributors

We gratefully acknowledge the following core contributors to N3C: Adam B. Wilcox, Adam M. Lee, Alexis Graves, Alfred (Jerrod) Anzalone, Amin Manna, Amit Saha, Amy Olex, Andrea Zhou, Andrew E. Williams, Andrew M. Southerland, Andrew T. Girvin, Anita Walden, Anjali Sharathkumar, Benjamin Amor, Benjamin Bates, Brian Hendricks, Brijesh Patel, G. Caleb Alexander, Carolyn T. Bramante, Cavin Ward-Caviness, Charisse Madlock-Brown, Christine Suver, Christopher G. Chute, Christopher Dillon, Chunlei Wu, Clare Schmitt, Cliff Takemoto, Dan Housman, Davera Gabriel, David A. Eichmann, Diego Mazzotti, Donald E. Brown, Eilis Boudreau, Elaine L. Hill, Emily Carlson Marti, Emily R. Pfaff, Evan French, Farrukh M Koraishy, Federico Mariona, Fred Prior, George Sokos, Greg Martin, Harold P. Lehmann, Heidi Spratt, Hemalkumar B. Mehta, J.W. Awori Hayanga, Jami Pincavitch, Jaylyn Clark, Jeremy Richard Harper, Jessica Yasmine Islam, Jin Ge, Joel Gagnier, Johanna J. Loomba, John B. Buse, Jomol Mathew, Joni L. Rutter, Julie A. McMurry, Justin Guinney, Justin Starren, Karen Crowley, Katie Rebecca Bradwell, Kellie M. Walters, Ken Wilkins, Kenneth R. Gersing, Kenrick Cato, Kimberly Murray, Kristin Kostka, Lavance Northington, Lee Pyles, Lesley Cottrell, Lili M. Portilla, Mariam Deacy, Mark M. Bissell, Marshall Clark, Mary Emmett, Matvey B. Palchuk, Melissa A. Haendel, Meredith Adams, Meredith Temple-O’Connor, Michael G. Kurilla, Michele Morris, Nasia Safdar, Nicole Garbarini, Noha Sharafeldin, Ofer Sadan, Patricia A. Francis, Penny Wung Burgoon, Philip R.O. Payne, Randeep Jawa, Rebecca Erwin-Cohen, Rena C. Patel, Richard A. Moffitt, Richard L. Zhu, Rishikesan Kamaleswaran, Robert Hurley, Robert T. Miller, Saiju Pyarajan, Sam G. Michael, Samuel Bozzette, Sandeep K. Mallipattu, Satyanarayana Vedula, Scott Chapman, Shawn T. O’Neil, Soko Setoguchi, Stephanie S. Hong, Steven G. Johnson, Tellen D. Bennett, Tiffany J. Callahan, Umit Topaloglu, Valery Gordon, Vignesh Subbian, Warren A. Kibbe, Wenndy Hernandez, Will Beasley, Will Cooper, William Hillegass, Xiaohan Tanner Zhang. Details of contributions available at covid.cd2h.org/core-contributors

Note: Google Scholar’s indexing accuracy can be compromised due to factors like language mismatches in text and metadata, incorrect author name formatting, and errors in automated bibliographic data extraction. Indexing may also take 6–8 weeks post-publication.

#### Data Partners with Released Data

The following institutions whose data is released or pending:

Available: Advocate Health Care Network — UL1TR002389: The Institute for Translational Medicine (ITM) • Aurora Health Care Inc — UL1TR002373: Wisconsin Network For Health Research • Boston University Medical Campus — UL1TR001430: Boston University Clinical and Translational Science Institute • Brown University — U54GM115677: Advance Clinical Translational Research (Advance-CTR) • Carilion Clinic — UL1TR003015: iTHRIV Integrated Translational health Research Institute of Virginia • Case Western Reserve University — UL1TR002548: The Clinical & Translational Science Collaborative of Cleveland (CTSC) • Charleston Area Medical Center — U54GM104942: West Virginia Clinical and Translational Science Institute (WVCTSI) • Children’s Hospital Colorado — UL1TR002535: Colorado Clinical and Translational Sciences Institute • Columbia University Irving Medical Center — UL1TR001873: Irving Institute for Clinical and Translational Research • Dartmouth College — None (Voluntary) Duke University — UL1TR002553: Duke Clinical and Translational Science Institute • George Washington Children’s Research Institute — UL1TR001876: Clinical and Translational Science Institute at Children’s National (CTSA-CN) • George Washington University — UL1TR001876: Clinical and Translational Science Institute at Children’s National (CTSA-CN) • Harvard Medical School — UL1TR002541: Harvard Catalyst • Indiana University School of Medicine — UL1TR002529: Indiana Clinical and Translational Science Institute • Johns Hopkins University — UL1TR003098: Johns Hopkins Institute for Clinical and Translational Research • Louisiana Public Health Institute — None (Voluntary) • Loyola Medicine — Loyola University Medical Center • Loyola University Medical Center — UL1TR002389: The Institute for Translational Medicine (ITM) • Maine Medical Center — U54GM115516: Northern New England Clinical & Translational Research (NNE-CTR) Network • Mary Hitchcock Memorial Hospital & Dartmouth Hitchcock Clinic — None (Voluntary) • Massachusetts General Brigham UL1TR002541: Harvard Catalyst • Mayo Clinic Rochester — UL1TR002377: Mayo Clinic Center for Clinical and Translational Science (CCaTS) • Medical University of South Carolina — UL1TR001450: South Carolina Clinical & Translational Research Institute (SCTR) • MITRE Corporation — None (Voluntary) • Montefiore Medical Center — UL1TR002556: Institute for Clinical and Translational Research at Einstein and Montefiore • Nemours — U54GM104941: Delaware CTR ACCEL Program • NorthShore University HealthSystem — UL1TR002389: The Institute for Translational Medicine (ITM) • Northwestern University at Chicago — UL1TR001422: Northwestern University Clinical and Translational Science Institute (NUCATS) • OCHIN — INV-018455: Bill and Melinda Gates Foundation grant to Sage Bionetworks • Oregon Health & Science University — UL1TR002369: Oregon Clinical and Translational Research Institute • Penn State Health Milton S. Hershey Medical Center — UL1TR002014: Penn State Clinical and Translational Science Institute • Rush University Medical Center — UL1TR002389: The Institute for Translational Medicine (ITM) • Rutgers, The State University of New Jersey — UL1TR003017: New Jersey Alliance for Clinical and Translational Science • Stony Brook University — U24TR002306 • The Alliance at the University of Puerto Rico, Medical Sciences Campus — U54GM133807: Hispanic Alliance for Clinical and Translational Research (The Alliance) • The Ohio State University — UL1TR002733: Center for Clinical and Translational Science • The State University of New York at Buffalo — UL1TR001412: Clinical and Translational Science Institute • The University of Chicago — UL1TR002389: The Institute for Translational Medicine (ITM) • The University of Iowa — UL1TR002537: Institute for Clinical and Translational Science • The University of Miami Leonard M. Miller School of Medicine — UL1TR002736: University of Miami Clinical and Translational Science Institute • The University of Michigan at Ann Arbor — UL1TR002240: Michigan Institute for Clinical and Health Research • The University of Texas Health Science Center at Houston — UL1TR003167: Center for Clinical and Translational Sciences (CCTS) • The University of Texas Medical Branch at Galveston — UL1TR001439: The Institute for Translational Sciences • The University of Utah — UL1TR002538: Uhealth Center for Clinical and Translational Science • Tufts Medical Center — UL1TR002544: Tufts Clinical and Translational Science Institute • Tulane University — UL1TR003096: Center for Clinical and Translational Science • The Queens Medical Center — None (Voluntary) • University Medical Center New Orleans — U54GM104940: Louisiana Clinical and Translational Science (LA CaTS) Center • University of Alabama at Birmingham — UL1TR003096: Center for Clinical and Translational Science • University of Arkansas for Medical Sciences — UL1TR003107: UAMS Translational Research Institute • University of Cincinnati — UL1TR001425: Center for Clinical and Translational Science and Training • University of Colorado Denver, Anschutz Medical Campus — UL1TR002535: Colorado Clinical and Translational Sciences Institute • University of Illinois at Chicago — UL1TR002003: UIC Center for Clinical and Translational Science • University of Kansas Medical Center — UL1TR002366: Frontiers: University of Kansas Clinical and Translational Science Institute • University of Kentucky — UL1TR001998: UK Center for Clinical and Translational Science • University of Massachusetts Medical School Worcester — UL1TR001453: The UMass Center for Clinical and Translational Science (UMCCTS) • University Medical Center of Southern Nevada — None (voluntary) • University of Minnesota — UL1TR002494: Clinical and Translational Science Institute • University of Mississippi Medical Center — U54GM115428: Mississippi Center for Clinical and Translational Research (CCTR) • University of Nebraska Medical Center — U54GM115458: Great Plains IDeA-Clinical & Translational Research • University of North Carolina at Chapel Hill — UL1TR002489: North Carolina Translational and Clinical Science Institute • University of Oklahoma Health Sciences Center — U54GM104938: Oklahoma Clinical and Translational Science Institute (OCTSI) • University of Pittsburgh — UL1TR001857: The Clinical and Translational Science Institute (CTSI) • University of Pennsylvania — UL1TR001878: Institute for Translational Medicine and Therapeutics • University of Rochester — UL1TR002001: UR Clinical & Translational Science Institute • University of Southern California — UL1TR001855: The Southern California Clinical and Translational Science Institute (SC CTSI) • University of Vermont — U54GM115516: Northern New England Clinical & Translational Research (NNE-CTR) Network • University of Virginia — UL1TR003015: iTHRIV Integrated Translational health Research Institute of Virginia • University of Washington — UL1TR002319: Institute of Translational Health Sciences • University of Wisconsin-Madison — UL1TR002373: UW Institute for Clinical and Translational Research • Vanderbilt University Medical Center — UL1TR002243: Vanderbilt Institute for Clinical and Translational Research • Virginia Commonwealth University — UL1TR002649: C. Kenneth and Dianne Wright Center for Clinical and Translational Research • Wake Forest University Health Sciences — UL1TR001420: Wake Forest Clinical and Translational Science Institute • Washington University in St. Louis — UL1TR002345: Institute of Clinical and Translational Sciences • Weill Medical College of Cornell University — UL1TR002384: Weill Cornell Medicine Clinical and Translational Science Center • West Virginia University — U54GM104942: West Virginia Clinical and Translational Science Institute (WVCTSI)  Submitted: Icahn School of Medicine at Mount Sinai — UL1TR001433: ConduITS Institute for Translational Sciences • The University of Texas Health Science Center at Tyler — UL1TR003167: Center for Clinical and Translational Sciences (CCTS) • University of California, Davis — UL1TR001860: UCDavis Health Clinical and Translational Science Center • University of California, Irvine — UL1TR001414: The UC Irvine Institute for Clinical and Translational Science (ICTS) • University of California, Los Angeles — UL1TR001881: UCLA Clinical Translational Science Institute • University of California, San Diego — UL1TR001442: Altman Clinical and Translational Research Institute • University of California, San Francisco — UL1TR001872: UCSF Clinical and Translational Science Institute  NYU Langone Health Clinical Science Core, Data Resource Core, and PASC Biorepository Core — OTA-21-015A: Post-Acute Sequelae of SARS-CoV-2 Infection Initiative (RECOVER)  Pending: Arkansas Children’s Hospital — UL1TR003107: UAMS Translational Research Institute • Baylor College of Medicine — None (Voluntary) • Children’s Hospital of Philadelphia — UL1TR001878: Institute for Translational Medicine and Therapeutics • Cincinnati Children’s Hospital Medical Center — UL1TR001425: Center for Clinical and Translational Science and Training • Emory University — UL1TR002378: Georgia Clinical and Translational Science Alliance • HonorHealth — None (Voluntary) • Loyola University Chicago — UL1TR002389: The Institute for Translational Medicine (ITM) • Medical College of Wisconsin — UL1TR001436: Clinical and Translational Science Institute of Southeast Wisconsin • MedStar Health Research Institute — None (Voluntary) • Georgetown University — UL1TR001409: The Georgetown-Howard Universities Center for Clinical and Translational Science (GHUCCTS) • MetroHealth — None (Voluntary) • Montana State University — U54GM115371: American Indian/Alaska Native CTR • NYU Langone Medical Center — UL1TR001445: Langone Health’s Clinical and Translational Science Institute • Ochsner Medical Center — U54GM104940: Louisiana Clinical and Translational Science (LA CaTS) Center • Regenstrief Institute — UL1TR002529: Indiana Clinical and Translational Science Institute • Sanford Research — None (Voluntary) • Stanford University — UL1TR003142: Spectrum: The Stanford Center for Clinical and Translational Research and Education • The Rockefeller University — UL1TR001866: Center for Clinical and Translational Science • The Scripps Research Institute — UL1TR002550: Scripps Research Translational Institute • University of Florida — UL1TR001427: UF Clinical and Translational Science Institute • University of New Mexico Health Sciences Center — UL1TR001449: University of New Mexico Clinical and Translational Science Center • University of Texas Health Science Center at San Antonio — UL1TR002645: Institute for Integration of Medicine and Science • Yale New Haven Hospital — UL1TR001863: Yale Center for Clinical Investigation

