## Supplementary File for "Impact of Preexisting Rare Diseases on COVID-19 Severity, Reinfection, and Long COVID, and the Modifying Effects of Vaccination and Antiviral Therapy: A Retrospective Study from the N3C Data Enclave"

**Univariate & multivariate models adjusted for demographics and comorbidities (Table 2/S1)**

*Univariate model: (Severity ~ Rare disease(i)),*
*Multivariate model: (Severity ~~ Rare disease(i)+ Demographic + Comorbidities); i = 1,...,18.*

**A Multivariable Model Evaluating the Impact of Vaccination and Antiviral Therapy on COVID-19 Severity by Rare Disease Status (*Table3/S3*)**
{Treatment (vaccination and/or antiviral treatment) vs control}

*Severity ~ treatment + demographic + comorbidities*

**A Multivariable Model Evaluating the Impact of Vaccination and Antiviral Therapy on Long COVID by Rare Disease Status (*Table4*)**
{Treatment (vaccination and/or antiviral treatment) vs control}

*Long COVID ~ treatment + demographic + comorbidities*

**A Multivariable Model Evaluating the Impact of Vaccination on COVID reinfection by Rare Disease Status (*Table5*)**
{Treatment (vaccination vs control)}

*COVID Reinfection ~ treatment + demographic + comorbidities*

**Figure S1:** Statistical models evaluated among COVID-19 patients in the N3C to assess associations between preexisting rare diseases (RDs) and three outcomes: COVID-19 severity, long COVID, and reinfection. Severity outcomes include hospitalization (Yes/No) and life-threatening illness (Yes/No). Similar binary definitions are used for long COVID and reinfection outcomes. Models are adjusted for demographic covariates and preexisting comorbidities (see Methods).

### Table S1: Univariate logistic regression models of severity outcomes.

| | Life-threatening<br>Yes: 113,510<br>No: 4,712,095 | | | Hospitalized<br>Yes: 254,728<br>No: 4,570,877 | |
|---|---|---|---|---|---|
| | **Univariate OR (95% CI)** | ***P* value** | | **Univariate OR (95% CI)** | ***P* value** |
| Rare neoplastic disease | 5.12 (4.95 - 5.31) | <0.001 | Rare endocrine disease | 3.27 (3.0 - 3.56) | <0.001 |
| Rare respiratory disease | 4.69 (4.52 - 4.87) | <0.001 | Rare respiratory disease | 3.2 (3.1 - 3.3) | <0.001 |
| Rare endocrine disease | 3.42 (3.04 - 3.85) | <0.001 | Rare hematologic disease | 3.06 (2.97 - 3.15) | <0.001 |
| Rare gastroenterologic disease | 3.37 (3.11 - 3.66) | <0.001 | Rare neoplastic disease | 2.66 (2.58 - 2.75) | <0.001 |
| Rare hepatic disease | 3.18 (2.92 - 3.46) | <0.001 | Rare gastroenterologic disease | 2.56 (2.4 - 2.73) | <0.001 |
| Rare hematologic disease | 2.94 (2.82 - 3.07) | <0.001 | Rare bone diseases | 2.36 (2.15 - 2.6) | <0.001 |
| Rare bone diseases | 2.92 (2.58 - 3.31) | <0.001 | Rare renal disease | 2.31 (2.07 - 2.58) | <0.001 |
| Rare infectious disease | 2.78 (2.62 - 2.95) | <0.001 | Rare ophthalmic disorder | 2.3 (2.16 - 2.45) | <0.001 |
| Rare ophthalmic disorder | 2.57 (2.36 - 2.8) | <0.001 | Rare hepatic disease | 2.28 (2.13 - 2.44) | <0.001 |
| Rare systemic or rheumatologic disease | 2.56 (2.46 - 2.66) | <0.001 | Rare systemic or rheumatologic disease | 2.19 (2.13 - 2.26) | <0.001 |
| Rare cardiac diseases | 2.51 (2.17 - 2.9) | <0.001 | Rare infectious disease | 2.16 (2.07 - 2.27) | <0.001 |
| Rare neurologic disease | 2.39 (2.31 - 2.46) | <0.001 | Rare cardiac diseases | 2.14 (1.92 - 2.39) | <0.001 |
| Rare immune disease | 2.24 (2.04 - 2.45) | <0.001 | Rare neurologic disease | 2.11 (2.06 - 2.16) | <0.001 |
| Rare renal disease | 1.97 (1.67 - 2.33) | <0.001 | Rare developmental defect during embryogenesis | 2.04 (1.96 - 2.13) | <0.001 |
| Rare skin disease | 1.93 (1.78 - 2.1) | <0.001 | Rare immune disease | 1.81 (1.69 - 1.94) | <0.001 |
| Rare developmental defect during embryogenesis | 1.75 (1.65 - 1.87) | <0.001 | Rare otorhinolaryngologic disease | 1.72 (1.56 - 1.9) | <0.001 |
| Rare otorhinolaryngologic disease | 1.67 (1.44 - 1.93) | <0.001 | Rare inborn errors of metabolism | 1.57 (1.43 - 1.72) | <0.001 |
| Rare inborn errors of metabolism | 1.45 (1.26 - 1.67) | <0.001 | Rare skin disease | 1.52 (1.43 - 1.62) | <0.001 |

**Table S2: Characteristics of COVID-19 subcohort of patients with reliable treatment documentation (December 23rd, 2021, to Jan 4th, 2024, n = 799,662) for assessing the impact of vaccination and antiviral treatment.**

| | All patients<br>(N = 799,662) | Rare Disease (No)<br>(N = 733,641) | Rare disease (Yes)<br>(N = 66,021) |
|---|---|---|---|
| **COVID-19 prevention/intervention** | | | |
| Antiviral treatment, n (%) | 42,023 (5.3%) | 38,385 (5.2%) | 3,638 (5.5%) |
| Vaccination status, n (%) | 321,834 (40%) | 294,607 (40%) | 27,227 (41%) |
| Antiviral & vaccination, n (%) | 129,727 (16%) | 117,079 (16%) | 12,648 (19%) |
| Control, n (%) | 306,078 (38%) | 283,570 (39%) | 22,508 (34%) |
| **COVID-19 related outcomes** | | | |
| Life-threatening, n (%) | 16,712 (2.1%) | 13,443 (1.8%) | 3,269 (5.0%) |
| Hospitalized, n (%) | 39,464 (4.9%) | 33,161 (4.5%) | 6,303 (9.5%) |
| Long COVID, n (%) | 46,494 (5.8%) | 39,263 (5.4%) | 7,231 (11%) |
| **Demographic** | | | |
| Age, n (%) | | | |
| Age (1-20) | 93,389 (12%) | 88,385 (12%) | 5,004 (7.6%) |
| Age (20-40) | 192,475 (24%) | 180,839 (25%) | 11,636 (18%) |
| Age (40-65) | 298,635 (37%) | 274,026 (37%) | 24,609 (37%) |
| Age (>65) | 215,163 (27%) | 190,391 (26%) | 24,772 (38%) |
| BMI, n (%) | | | |
| Obese | 437,153 (55%) | 400,396 (55%) | 36,757 (56%) |
| Over weight | 197,732 (25%) | 181,155 (25%) | 16,577 (25%) |
| Normal | 130,938 (16%) | 120,544 (16%) | 10,394 (16%) |
| Under weight | 33,839 (4.2%) | 31,546 (4.3%) | 2,293 (3.5%) |
| Sex, n(%) | | | |
| Female | 493,441 (62%) | 451,950 (62%) | 41,491 (63%) |
| Male | 306,221 (38%) | 281,691 (38%) | 24,530 (37%) |
| Race, n(%) | | | |
| Asian | 41,367 (5.2%) | 38,846 (5.3%) | 2,521 (3.8%) |
| Black or African American | 105,988 (13%) | 96,045 (13%) | 9,943 (15%) |
| Missing/Unknown/Other | 93,058 (12%) | 87,287 (12%) | 5,771 (8.7%) |
| White | 559,249 (70%) | 511,463 (70%) | 47,786 (72%) |
| Ethinicity, n(%) | | | |
| Hispanic or Latino | 99,047 (12%) | 94,211 (13%) | 4,836 (7.3%) |
| Not hispanic or Latino | 670,302 (84%) | 610,705 (83%) | 59,597 (90%) |
| Missing/Unknown | 30,313 (3.8%) | 28,725 (3.9%) | 1,588 (2.4%) |
| Smoking status, n(%) | | | |
| Current or former smoker | 138,643 (17%) | 125,550 (17%) | 13,093 (20%) |
| Non-smoker | 661,019 (83%) | 608,091 (83%) | 52,928 (80%) |
| **Comorbidities, n(%)** | | | |
| Cancer | 92,773 (12%) | 74,629 (10%) | 18,144 (27%) |
| Cardiomyopathies | 27,141 (3.4%) | 21,661 (3.0%) | 5,480 (8.3%) |
| Cerebro vascular disease | 45,575 (5.7%) | 36,958 (5.0%) | 8,617 (13%) |
| Chronic lung disease | 179,477 (22%) | 152,497 (21%) | 26,980 (41%) |
| Coronary artery disease | 79,411 (9.9%) | 66,780 (9.1%) | 12,631 (19%) |
| Dementia before | 20,088 (2.5%) | 17,073 (2.3%) | 3,015 (4.6%) |
| Depression | 182,887 (23%) | 160,081 (22%) | 22,806 (35%) |
| Diabetes | 142,647 (18%) | 125,383 (17%) | 17,264 (26%) |
| Heart failure | 60,220 (7.5%) | 48,172 (6.6%) | 12,048 (18%) |
| HIV infection | 6,144 (0.8%) | 5,208 (0.7%) | 936 (1.4%) |
| Hypertension | 310,976 (39%) | 274,323 (37%) | 36,653 (56%) |
| Kidney disease | 81,261 (10%) | 66,679 (9.1%) | 14,582 (22%) |
| Liver disease | 61,810 (7.7%) | 50,261 (6.9%) | 11,549 (17%) |
| Myocardial infraction | 34,945 (4.4%) | 28,716 (3.9%) | 6,229 (9.4%) |
| Peripheral vascular disease | 38,007 (4.8%) | 31,268 (4.3%) | 6,739 (10%) |
| Rheumatologic disease | 64,127 (8.0%) | 49,659 (6.8%) | 14,468 (22%) |

### Table S3: Impact of vaccination and antiviral treatment exposures on hospitalization in patients with and without preexising RDs (multivariate logistic regression models are adjusted for demographics and comorbidities).

| | Patients with Preexisting Rare Disease | | | | | | Patients without Preexisting Rare Disease | | | | | |
|---|---|---|---|---|---|---|---|---|---|---|---|---|
| | Vaccination vs Control | | Antiviral treatment vs Control | | Vaccination + antiviral treatment vs Control | | Vaccination vs Control | | Antiviral treatment vs Control | | Vaccination + antiviral treatment vs Control | |
| | Hospitalized condition Yes: 5,888 No: 43,847 | | Hospitalized condition Yes: 3,192 No: 22,954 | | Hospitalized condition Yes: 6,303 No: 59,718 | | Hospitalized condition Yes: 30,958 No: 547,219 | | Hospitalized condition Yes: 19,727 No: 302,228 | | Hospitalized condition Yes: 33,161 No: 700,480 | |
| | **OR (95% CI)** | **P value** | **OR (95% CI)** | **P value** | **OR (95% CI)** | **P value** | **OR (95% CI)** | **P value** | **OR (95% CI)** | **P value** | **OR (95% CI)** | **P value** |
| **)-19 prevention/intervention** | | | | | | | | | | | | |
| :ol | 1 [Reference] | | 1 [Reference] | | 1 [Reference] | | 1 [Reference] | | 1 [Reference] | | 1 [Reference] | |
| Vaccination status | **0.62 (0.59 - 0.66)** | **< 0.001** | | | | | **0.45 (0.44 - 0.47)** | **< 0.001** | | | | |
| Antiviral treatment | | | **0.31 (0.26 - 0.37)** | **< 0.001** | | | | | **0.34 (0.31 - 0.36)** | **< 0.001** | | |
| Vaccination & Antiviral | | | | | **0.11 (0.1 - 0.13)** | **< 0.001** | | | | | **0.09 (0.09 - 0.1)** | **< 0.001** |
| **Demographic** | | | | | | | | | | | | |
| **Age** | | | | | | | | | | | | |
| Age (20-40) | 1 [Reference] | | 1 [Reference] | | 1 [Reference] | | 1 [Reference] | | 1 [Reference] | | 1 [Reference] | |
| Age (1-20) | 1.05 (0.9 - 1.24) | 0.540 | 0.89 (0.75 - 1.05) | 0.167 | 0.92 (0.77 - 1.09) | 0.312 | 0.48 (0.45 - 0.52) | < 0.001 | 0.5 (0.47 - 0.54) | < 0.001 | 0.48 (0.44 - 0.52) | < 0.001 |
| Age (40-65) | 0.93 (0.84 - 1.02) | 0.121 | 1.04 (0.92 - 1.17) | 0.498 | 1.01 (0.9 - 1.14) | 0.811 | 1.02 (0.99 - 1.07) | 0.225 | 1.02 (0.97 - 1.06) | 0.470 | 1.01 (0.96 - 1.05) | 0.808 |
| Age (>65) | 1.22 (1.1 - 1.36) | < 0.001 | 1.27 (1.11 - 1.46) | 0.001 | 1.27 (1.11 - 1.45) | 0.001 | 1.85 (1.77 - 1.94) | < 0.001 | 1.83 (1.73 - 1.93) | < 0.001 | 1.78 (1.69 - 1.89) | < 0.001 |
| **BMI** | | | | | | | | | | | | |
| Normal | 1 [Reference] | | 1 [Reference] | | 1 [Reference] | | 1 [Reference] | | 1 [Reference] | | 1 [Reference] | |
| Obese | 0.75 (0.69 - 0.82) | < 0.001 | 0.72 (0.64 - 0.81) | < 0.001 | 0.71 (0.63 - 0.8) | < 0.001 | 0.72 (0.7 - 0.75) | < 0.001 | 0.76 (0.72 - 0.79) | < 0.001 | 0.71 (0.68 - 0.75) | < 0.001 |
| Over weight | 0.77 (0.7 - 0.85) | < 0.001 | 0.8 (0.7 - 0.91) | 0.001 | 0.77 (0.68 - 0.88) | < 0.001 | 0.83 (0.8 - 0.87) | < 0.001 | 0.87 (0.82 - 0.91) | < 0.001 | 0.81 (0.77 - 0.86) | < 0.001 |
| Under weight | 0.79 (0.63 - 0.98) | 0.030 | 0.76 (0.61 - 0.94) | 0.011 | 0.8 (0.64 - 1) | 0.050 | 0.82 (0.74 - 0.91) | < 0.001 | 0.7 (0.63 - 0.78) | < 0.001 | 0.71 (0.64 - 0.79) | < 0.001 |
| **Sex** | | | | | | | | | | | | |
| Female | 1 [Reference] | | 1 [Reference] | | 1 [Reference] | | 1 [Reference] | | 1 [Reference] | | 1 [Reference] | |
| Male | 1.3 (1.22 - 1.38) | < 0.001 | 1.32 (1.21 - 1.43) | < 0.001 | 1.29 (1.19 - 1.41) | < 0.001 | 1.17 (1.14 - 1.2) | < 0.001 | 1.13 (1.09 - 1.16) | < 0.001 | 1.16 (1.12 - 1.2) | < 0.001 |
| **Race** | | | | | | | | | | | | |
| White | 1 [Reference] | | 1 [Reference] | | 1 [Reference] | | 1 [Reference] | | 1 [Reference] | | 1 [Reference] | |
| Asian | 1.01 (0.86 - 1.2) | 0.887 | 1.01 (0.81 - 1.26) | 0.937 | 0.99 (0.78 - 1.24) | 0.905 | 0.9 (0.85 - 0.96) | 0.002 | 0.91 (0.84 - 0.98) | 0.016 | 0.93 (0.85 - 1.01) | 0.077 |
| Black or African American | 1.67 (1.54 - 1.8) | < 0.001 | 1.58 (1.42 - 1.76) | < 0.001 | 1.56 (1.4 - 1.74) | < 0.001 | 1.36 (1.31 - 1.41) | < 0.001 | 1.4 (1.34 - 1.47) | < 0.001 | 1.39 (1.33 - 1.45) | < 0.001 |
| Missing/Unknown/Other | 1.2 (1.06 - 1.35) | 0.004 | 1.17 (1 - 1.35) | 0.043 | 1.21 (1.04 - 1.41) | 0.014 | 1.17 (1.11 - 1.22) | < 0.001 | 1.13 (1.07 - 1.2) | < 0.001 | 1.15 (1.09 - 1.22) | < 0.001 |
| **Ethinicity** | | | | | | | | | | | | |
| Not hispanic or Latino | 1 [Reference] | | 1 [Reference] | | 1 [Reference] | | 1 [Reference] | | 1 [Reference] | | 1 [Reference] | |
| Hispanic or Latino | 1.19 (1.04 - 1.36) | 0.009 | 1.25 (1.07 - 1.47) | 0.005 | 1.2 (1.02 - 1.42) | 0.029 | 0.8 (0.76 - 0.84) | < 0.001 | 0.9 (0.85 - 0.95) | < 0.001 | 0.9 (0.84 - 0.96) | 0.001 |
| Missing/Unknown | 0.45 (0.34 - 0.6) | < 0.001 | 0.45 (0.31 - 0.64) | < 0.001 | 0.42 (0.29 - 0.61) | < 0.001 | 0.46 (0.42 - 0.51) | < 0.001 | 0.51 (0.45 - 0.57) | < 0.001 | 0.47 (0.41 - 0.53) | < 0.001 |
| **Smoking status** | | | | | | | | | | | | |
| Non-smoker | 1 [Reference] | | 1 [Reference] | | 1 [Reference] | | 1 [Reference] | | 1 [Reference] | | 1 [Reference] | |
| Current or former smoker | 1.42 (1.32 - 1.53) | < 0.001 | 1.15 (1.04 - 1.28) | 0.005 | 1.18 (1.07 - 1.31) | 0.001 | 1.39 (1.35 - 1.44) | < 0.001 | 1.18 (1.14 - 1.23) | < 0.001 | 1.19 (1.14 - 1.24) | < 0.001 |
| **Comorbidities** | | | | | | | | | | | | |
| :er | 1.34 (1.25 - 1.43) | < 0.001 | 1.31 (1.2 - 1.45) | < 0.001 | 1.35 (1.22 - 1.48) | < 0.001 | 1.31 (1.26 - 1.35) | < 0.001 | 1.29 (1.23 - 1.35) | < 0.001 | 1.29 (1.23 - 1.36) | < 0.001 |
| liomyophaties | 0.97 (0.87 - 1.07) | 0.534 | 0.87 (0.75 - 1.01) | 0.069 | 0.89 (0.77 - 1.03) | 0.126 | 1.02 (0.96 - 1.08) | 0.520 | 1 (0.93 - 1.08) | 0.897 | 1.01 (0.93 - 1.08) | 0.873 |
| :bro vascular disease | 1.26 (1.17 - 1.37) | < 0.001 | 1.28 (1.14 - 1.44) | < 0.001 | 1.3 (1.16 - 1.46) | < 0.001 | 1.32 (1.26 - 1.37) | < 0.001 | 1.32 (1.25 - 1.39) | < 0.001 | 1.34 (1.26 - 1.41) | < 0.001 |
| )nic lung disease | 1.37 (1.29 - 1.46) | < 0.001 | 1.43 (1.31 - 1.55) | < 0.001 | 1.4 (1.28 - 1.52) | < 0.001 | 1.46 (1.42 - 1.51) | < 0.001 | 1.44 (1.39 - 1.5) | < 0.001 | 1.49 (1.44 - 1.55) | < 0.001 |
| )nary artery disease | 1.03 (0.94 - 1.12) | 0.567 | 0.97 (0.86 - 1.1) | 0.630 | 0.98 (0.87 - 1.1) | 0.711 | 1.03 (0.99 - 1.08) | 0.134 | 1 (0.95 - 1.06) | 0.923 | 1.01 (0.96 - 1.07) | 0.639 |
| ıentia before | 1.63 (1.45 - 1.82) | < 0.001 | 1.37 (1.16 - 1.61) | < 0.001 | 1.48 (1.25 - 1.74) | < 0.001 | 2.22 (2.11 - 2.33) | < 0.001 | 2.02 (1.89 - 2.16) | < 0.001 | 2.13 (1.99 - 2.28) | < 0.001 |
| :ession | 1.08 (1.01 - 1.15) | 0.026 | 1.11 (1.01 - 1.22) | 0.024 | 1.11 (1.01 - 1.21) | 0.029 | 1.11 (1.08 - 1.14) | < 0.001 | 1.13 (1.09 - 1.18) | 0.000 | 1.13 (1.08 - 1.18) | < 0.001 |
| etes | 1.25 (1.17 - 1.34) | < 0.001 | 1.29 (1.17 - 1.42) | < 0.001 | 1.27 (1.15 - 1.4) | < 0.001 | 1.39 (1.35 - 1.44) | < 0.001 | 1.4 (1.35 - 1.46) | < 0.001 | 1.41 (1.35 - 1.47) | < 0.001 |
| :t failure | 1.61 (1.48 - 1.76) | < 0.001 | 1.5 (1.33 - 1.7) | < 0.001 | 1.54 (1.37 - 1.74) | < 0.001 | 1.93 (1.85 - 2.02) | < 0.001 | 1.8 (1.7 - 1.91) | < 0.001 | 1.8 (1.7 - 1.91) | < 0.001 |
| infection | 1.01 (0.8 - 1.28) | 0.905 | 1.06 (0.8 - 1.42) | 0.678 | 1.09 (0.8 - 1.47) | 0.595 | 1.19 (1.03 - 1.37) | 0.015 | 1.32 (1.11 - 1.57) | 0.002 | 1.26 (1.05 - 1.51) | 0.013 |
| Hypertension | 1.41 (1.3 - 1.54) | < 0.001 | 1.47 (1.32 - 1.64) | < 0.001 | 1.49 (1.34 - 1.67) | < 0.001 | 1.62 (1.56 - 1.67) | < 0.001 | 1.59 (1.52 - 1.66) | < 0.001 | 1.58 (1.51 - 1.65) | < 0.001 |
| ıey disease | 1.64 (1.53 - 1.76) | < 0.001 | 1.61 (1.46 - 1.77) | < 0.001 | 1.62 (1.47 - 1.78) | < 2e-16 | 1.71 (1.65 - 1.77) | < 0.001 | 1.66 (1.59 - 1.74) | < 0.001 | 1.64 (1.57 - 1.72) | < 0.001 |
| : disease | 1.22 (1.14 - 1.32) | < 0.001 | 1.28 (1.16 - 1.42) | < 0.001 | 1.3 (1.17 - 1.44) | < 0.001 | 1.39 (1.34 - 1.45) | < 0.001 | 1.44 (1.36 - 1.51) | < 0.001 | 1.4 (1.32 - 1.48) | < 0.001 |
| cardial infraction | 1.25 (1.14 - 1.37) | < 0.001 | 1.3 (1.15 - 1.49) | < 0.001 | 1.33 (1.17 - 1.52) | < 0.001 | 1.55 (1.47 - 1.62) | < 0.001 | 1.56 (1.47 - 1.66) | < 0.001 | 1.54 (1.45 - 1.63) | < 0.001 |
| )heral vascular disease | 1.11 (1.01 - 1.21) | 0.025 | 1.04 (0.92 - 1.18) | 0.537 | 1.06 (0.94 - 1.21) | 0.346 | 1.06 (1.02 - 1.11) | 0.008 | 1.03 (0.96 - 1.09) | 0.429 | 1.05 (0.98 - 1.11) | 0.147 |
| ımatologic disease | 0.96 (0.9 - 1.03) | 0.277 | 0.85 (0.77 - 0.94) | 0.002 | 0.87 (0.78 - 0.96) | 0.006 | 1.03 (0.99 - 1.08) | 0.117 | 0.99 (0.93 - 1.04) | 0.611 | 1.02 (0.97 - 1.08) | 0.464 |
